# Flipped C-Terminal Ends of APOA1 Promote ABCA1-dependent Cholesterol Efflux by Small HDLs

**DOI:** 10.1101/2023.11.03.23297986

**Authors:** Yi He, Chiara Pavanello, Patrick M. Hutchins, Chongren Tang, Mohsen Pourmousa, Tomas Vaisar, Hyun D. Song, Richard W. Pastor, Alan T. Remaley, Ira J. Goldberg, Tina Costacou, W. Sean Davidson, Karin E. Bornfeldt, Laura Calabresi, Jere P. Segrest, Jay W. Heinecke

**Author notes:** **Correspondence**: Jay Heinecke, 850 Republican St, Box 358055, UW Medicine, Seattle, WA 98109 USA. **Abbreviations and acronyms**: ABCA1, ATP-binding cassette transporter A1; APOA1, apolipoprotein A1; APOB, apolipoprotein B; BHK, baby hamster kidney; calibrated IMA, calibrated ion mobility analysis; CEC, cholesterol efflux capacity; CVD, cardiovascular disease; EDC, 1-ethyl-3-(3-dimethylaminopropyl)carbodiimide hydrochloride; HDL-C, high-density lipoprotein cholesterol; HDL-P, HDL particle concentration determined by calibrated IMA; LCAT, lecithin–cholesterol acyltransferase; LDL-C, low-density lipoprotein cholesterol; L-HDL, large HDL; M-HDL, medium HDL; MS, mass spectrometry; r-HDL, reconstituted HDL; S-HDL, small HDL; XS-HDL, extra small HDL; MD, molecular dynamics.

## Abstract

**Background:** Cholesterol efflux capacity (CEC) predicts cardiovascular disease (CVD) independently of HDL cholesterol (HDL-C) levels. Isolated small HDL particles are potent promoters of macrophage CEC by the ABCA1 pathway, but the underlying mechanisms are unclear.

**Methods:** We used model system studies of reconstituted HDL and plasma from control and lecithin-cholesterol acyltransferase (LCAT)-deficient subjects to investigate the relationships among the sizes of HDL particles, the structure of APOA1 in the different particles, and the CECs of plasma and isolated HDLs.

**Results:** We quantified macrophage and ABCA1 CEC of four distinct sizes of reconstituted HDL (r-HDL). CEC increased as particle size decreased. MS/MS analysis of chemically crosslinked peptides and molecular dynamics simulations of APOA1 (HDL’s major protein) indicated that the mobility of that protein’s C-terminus was markedly higher and flipped off the surface in the smallest particles. To explore the physiological relevance of the model system studies, we isolated HDL from LCAT-deficient subjects, whose small HDLs–like r-HDLs–are discoidal and composed of APOA1, cholesterol, and phospholipid. Despite their very low plasma levels of HDL particles, these subjects had normal CEC. In both the LCAT-deficient subjects and control subjects, the CEC of isolated extra-small HDL (a mixture of extra-small and small HDL by calibrated ion mobility analysis) was 3–5-fold greater than that of the larger sizes of isolated HDL. Incubating LCAT-deficient plasma and control plasma with human LCAT converted extra-small and small HDL particles into larger particles, and it markedly inhibited CEC.

**Conclusions:** We present a mechanism for the enhanced CEC of small HDLs. In smaller particles, the C-termini of the two antiparallel molecules of APOA1 are flipped off the lipid surface of HDL. This extended conformation allows them to engage with ABCA1. In contrast, the C-termini of larger HDLs are unable to interact productively with ABCA1 because they form a helical bundle that strongly adheres to the lipid on the particle. Enhanced CEC, as seen with the smaller particles, predicts decreased CVD risk. Thus, extra-small and small HDLs may be key mediators and indicators of HDL’s cardioprotective effects.

**Clinical Perspective:**

- Using chemical crosslinking and molecular dynamics simulations, we showed that the C-termini of APOA1, HDL’s major protein, have increased mobility and conformational freedom in small HDL particles.
- The enhanced mobility of the C-termini of APOA1 in small HDLs allows the C-termini to ‘flip’ off a particle’s surface, activating ABCA1 thereby stimulating cholesterol removal from cells.
- Because of small HDLs’ vital role in cholesterol efflux, quantification of HDL-P (the size and concentration of HDL subspecies) might be a better metric for gauging cardiovascular disease risk than HDL-cholesterol levels.
- Therapeutic interventions that increase small HDL levels, with or without increasing HDL-cholesterol levels, may be cardioprotective.

## Introduction

The risk of cardiovascular disease (CVD) strongly and inversely associates with plasma levels of high-density lipoprotein cholesterol (HDL-C).^1^ However, pharmacological interventions that elevate HDL-C have failed to lower CVD risk in statin-treated subjects, suggesting that the association between HDL-C and CVD risk is indirect.^2^ It is therefore critical to identify new mechanisms that inversely link HDL to CVD risk and do not involve HDL-C.^3,4^

One proposed cardioprotective function of HDL is promotion of cholesterol efflux from lipid-laden macrophages, which play critical roles in all stages of atherogenesis.^5^ Two early steps in this pathway involve ATP-binding cassette transporters—ABCA1 and ABCG1. Initially, ABCA1 mediates cholesterol efflux from macrophages to lipid-poor apolipoproteins^6^ and small dense HDL.^7,8^ Lecithin-cholesterol acyltransferase (LCAT) then promotes HDL maturation by catalyzing the conversion of free cholesterol to cholesteryl esters, which are then transferred from the surface to the core, generating larger HDL particles.^9^ ABCG1 exports cellular cholesterol to larger HDL particles that deliver cholesterol to the liver for excretion in bile.^2^

Rothblat, Rader, and colleagues demonstrated that serum HDL (serum depleted of lipoproteins that contain apolipoprotein B [APOB]) promotes cholesterol efflux from cultured macrophages, thus mimicking the key early steps in reverse cholesterol transport from macrophages.^10,11^ The magnitude of cholesterol efflux to serum HDL, termed cholesterol efflux capacity (CEC), is largely independent of HDL-C.^10,11^ However, large clinical studies demonstrate that macrophage CEC and ABCA1-specific CEC of serum HDL strongly and negatively associate with prevalent and incident CVD.^12^ Importantly, CEC predicts CVD independently of HDL-C.^11–13^ These results suggest that CEC is a critical contributor to HDL’s proposed anti-atherogenic functions in humans.

Lecithin-cholesterol acyltransferase (LCAT) is widely regarded as an important driving force for mobilizing cholesterol from tissues to the liver for excretion. However, subjects with complete LCAT deficiency do not appear to be at increased risk for CVD^14^ despite having very low HDL-C levels. Animal models of atherosclerosis have yielded conflicting results on the impact of LCAT deficiency and overexpression.^15,16^ Serum from LCAT-deficient subjects exhibits elevated ABCA1-dependent cellular cholesterol efflux even though efflux via ABCG1 and SR-B1 is impaired, suggesting that cholesterol efflux by the ABCA1 pathway might explain why those subjects are not at high risk for CVD.^17^

CSL-112, a reconstituted HDL particle composed of human APOA1 and phosphatidylcholine, was designed to mimic small HDLs.^18^ The drug, which markedly enhances the ABCA1 CEC of human plasma, promotes the remodeling of HDL resulting in higher levels of small and lipid-poor APOA1 particles.^19^

Small HDLs account for most of serum HDL’s CEC activity.^7,8^ However, the underlying mechanisms are poorly understood. To explore potential mechanisms, we combined functional and structural studies of reconstituted HDL particles (r-HDL) with studies of control and LCAT-deficient subjects. Our studies reveal that the C-terminus of APOA1 in smaller HDLs becomes available to engage ABCA1, the first key step in cholesterol export from cells.

## Experimental Methods

### Generation of reconstituted HDL (r-HDL) particles

Discoidal r-HDL was prepared from recombinant human APOA1, 1-palmitoyl-oleoyl-phosphatidylcholine (POPC), and free cholesterol by cholate dialysis.^20–22^ The composition of the different size of particles are (APOA1:free cholesterol:POPC, mol/mol): r-HDL-80, 1.0:1.8:34; r-HDL-88, 1.0:2.9:52.7; r-HDL-96, 1.0:4.7:90.6; r-HDL-120, 1.0:4.7:140.^22^

### Calibrated ion mobility analysis (IMA)

The sizes of r-HDL and human HDL particles were quantified with a scanning mobility particle sizer spectrometer (TSI Inc., Shoreview, MN, model 3080N).^23–25^ The concentrations of r-HDL and HDL particles (mol/L) were determined using a calibration curve of glucose oxidase.^23^

### Chemical crosslinking of r-HDL

Reconstituted HDLs (r-HDLs) were crosslinked with 1-ethyl-(3-dimethylaminopropyl) carbodiimide hydrochloride (EDC) in phosphate-buffered saline (pH 6.5),^20,26^ and further fractionated by high-resolution size exclusion chromatography to isolate monomeric HDL particles. Details are provided in Supplemental Material.

### Proteolytic digestion and mass spectrometry analysis

Details are provided in Supplemental Material.

### Molecular dynamics simulations of r-HDL

Molecular dynamics (MD) trajectories of r-HDL-80 and r-HDL-90 were calculated using a combination of all-atom simulation, simulated tempering, and coarse-grained (CG) methods (Supplemental Material).^27–30^ Simulations of r-HDL-100 and r-HDL-120 (termed r-HDL-110) were reported previously.^27^ Because different preparations of the largest r-HDL particles range in size from 110–120 Å,^22^ we term these r-HDL-120 to be consistent with the size of the largest particles used here.

### HDL contact map analyses

Inter- and intramolecular contact maps between Cα atoms used a cutoff distance of 15.1 Å.^27^ A total of 2,084 and 1,042 frames from the last half of 20 µs and 10 µs simulations were used for the r-HDL-100 and r-HDL-120 particles, respectively.^27^ A total of 100 frames extracted from the last half of 200 µs CG simulations of r-HDL-80 and r-HDL-90 particles were converted to all-atom structures to develop the contact maps. Contact maps were plotted using Gnuplot version 5.2 (http://gnuplot.info).

### Cholesterol efflux capacity (CEC)

Macrophage cholesterol efflux capacity was assessed with J774 macrophages labeled with [^3^H]cholesterol and stimulated with a cAMP analog.^10^ Efflux via the ABCA1 pathway was measured with baby hamster kidney (BHK) cells that expressed mifepristone-inducible human ABCA1 and were labeled with [^3^H]cholesterol.^6^

### LCAT-deficient and control subjects

Twenty-four subjects; 4 carriers of two mutant LCAT alleles (termed LCAT-/- subjects), 6 carriers of 1 mutant LCAT allele (LCAT+/-), and 14 non-carriers (LCAT+/+) were from an Italian family study. The Italian LCAT deficient cohort includes related carriers (**Supplemental Table S1**). The study was approved by the institutional ethical committee.^31^ All subjects gave informed consent. Aliquots of serum from subjects who had fasted overnight were immediately frozen and stored at −80°C until analysis. Serum lipid levels and LCAT activity of the Italian cohort were determined as described.^31^

### Incubation of control and LCAT-deficient plasma with LCAT

Details are provided in Supplemental Material.^32^

### HDL Isolation

For functional studies, HDL was first isolated by ultracentrifugation (density 1.063–1.210 g/mL)^33^ from serum of control subjects (n=4) and LCAT-/- subjects (n=5) and then fractionated on a Superdex 200 Increase 10/300 GL column. Details are provided in Supplemental Material.

### Data availability

All data supporting the findings of this study are available in the article and/or its supplemental materials.

### Statistical Analyses

Statistical analyses were performed with STATA software version 12 (Stata Corp, College Park, TX) and with SAS v.9.4 (SAS Inc, Cary, NC, USA). Mixed effect models, considering family as a random effect, with Tukey-Kramer post-hoc tests were used to compare the means of three or more groups. One-way ANOVA was used to analyze laboratory experiments. Linear regression was used to investigate the correlation of serum HDL CEC with HDL particle concentration for each subspecies. Parametric or non-parametric analyses were based on the Shapiro-Wilk test for normality. The ratio t-test (GraphPad) was used for the analysis of plasma incubations with/without LCAT. The null hypothesis is that the average of the logarithms of the ratio of each pair is zero*. P*-values <0.05 were considered significant. Unless otherwise stated, values represent means ± standard deviations.

## Results

### Reconstituted small HDL particles promote macrophage CEC and ABCA1 CEC as effectively as lipid-free APOA1

We used reconstituted discoidal HDL (r-HDL) as a model system to investigate how particle size affects HDL’s ability to promote CEC.^20,22^ To quantify cholesterol efflux by macrophages and the ABCA1 pathway, we used validated model systems.^8,10,11^ r-HDLs were fractionated into 4 different sizes of particles, using high-resolution size exclusion chromatography.^20^ We term these particles r-HDL-80, r-HDL-88, r-HDL-96, and r-HDL-120 because their diameters are respectively 80 Å, 88 Å, 96 Å, and 110-120 Å as determined by calibrated IMA (**Fig. 1A**). These values are in excellent agreement with those previously determined by non-denaturing gradient gel electrophoresis and by quantification of the hydrodynamic Stokes’ diameters of the particles.^22^

**Figure 1.**
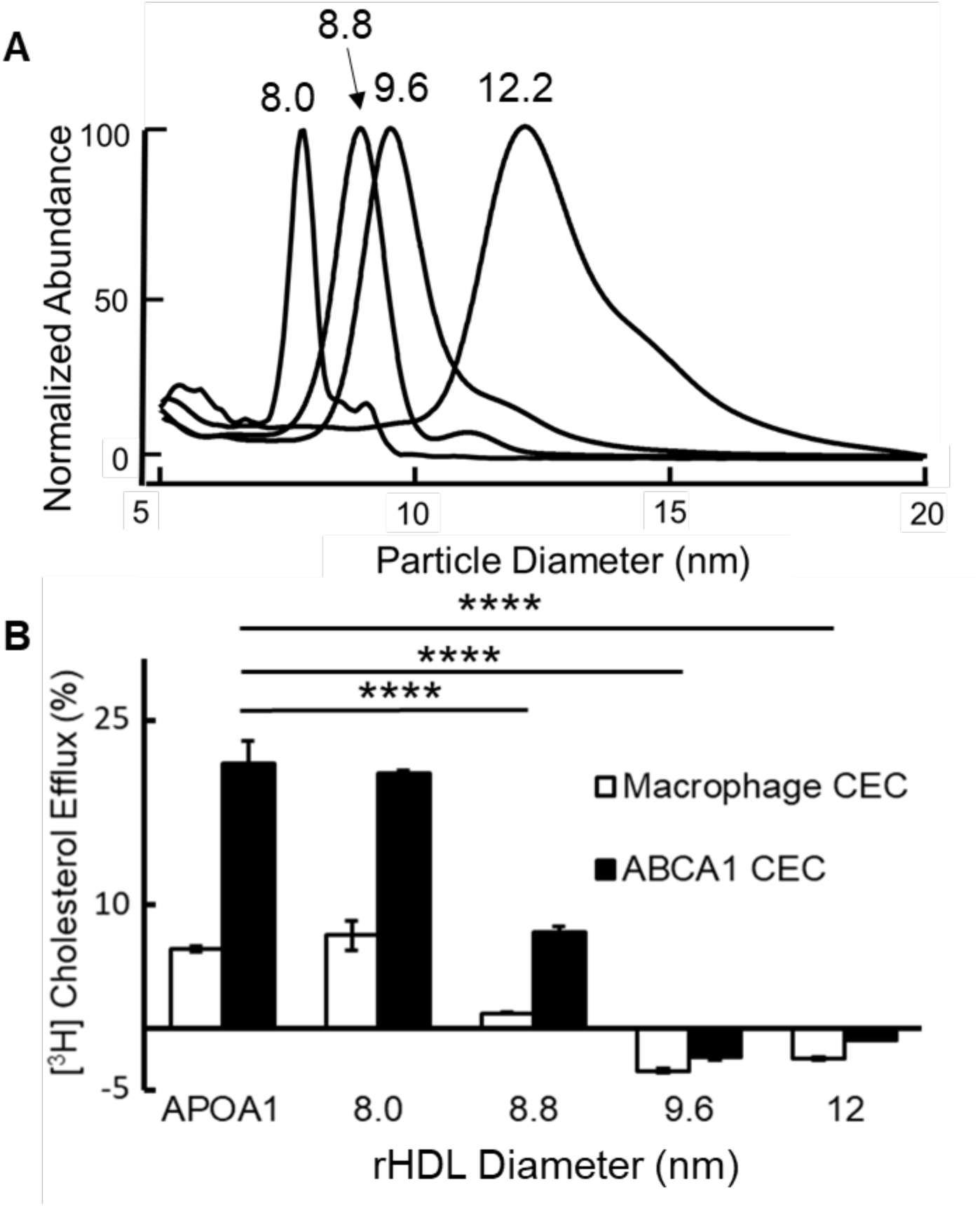
Calibrated IMA (A) and CEC (B) of reconstituted HDLs (r-HDLs) prepared by cholate dialysis and fractionation by high-resolution size exclusion chromatography. (**A**) Representative IMA profiles of each size of r-HDL. To facilitate comparison of the size distributions of the particles, the height of each r-HDL was set to 100%. The median sizes of the isolated particles were 8.0±0.2 nm, 8.8±0.1 nm, 9.6±0.1 nm, and 12.2±0.1 nm. (**B**) ABCA1-mediated cholesterol efflux capacity (CEC) using equimolar concentrations of each size of r-HDL. Macrophage CEC and ABCA1 CEC of serum HDL were quantified after a 4-h incubation with [^3^H]cholesterol-labeled J774 macrophages and BHK cells, without or with induction of ABCA1 expression with cAMP and mifepristone, respectively. Cholesterol efflux was calculated as the percentage of radiolabel in the medium of the cells divided by the total radioactivity of the medium and cells. CEC was quantified as the difference in cholesterol efflux of cells with and without induced expression of ABCA1. Results are representative of 5 independent experiments with replicate analyses. \*\*\*\**P*<0.001, one-way ANOVA with Tukey-Kramer post-tests.

Each particle population exhibited a symmetrical Gaussian-like distribution and was clearly distinguishable from the other sizes of particles by IMA (**Fig. 1A**). The r-HDL particles were similar in size to human XS-HDL, S-HDL, M-HDL, and L-HDL (see below).

At equimolar concentrations, the smallest r-HDL-80 particles were as effective as lipid-free APOA1 at promoting both macrophage CEC and ABCA1 CEC (**Fig. 1B**). The r-HDL-88 particles were less effective, and the two largest r-HDL particles (r-HDL-96 and r-HDL-120) failed to promote either macrophage CEC or ABCA1 CEC.

### Probing the structure of APOA1 in r-HDLs with MS/MS

In their double belt model for HDL, Segrest et al.^34^ proposed that 2 molecules of APOA1 form an anti-parallel helical bundle that encircles the edge of the discoidal HDL particle. The crystal structure of N-terminally truncated APOA1^35^ and chemical crosslinking studies of r-HDL^20,27,36^ and human HDL^20^ support this model.

In contrast to APOA1’s central region, which forms a stable helical bundle, the N-and C-terminal regions of APOA1 in HDL are more flexible and capable of assuming a variety of conformations.^22,27,36,37^ These regions of the protein are also important for promoting ABCA1-dependent cholesterol efflux.^36^ To investigate the structures of the different regions of APOA1 in the different sizes of r-HDL, we used EDC (1-ethyl-3-(3-dimethylaminopropyl)carbodiimide hydrochloride)^20^ to generate intramolecular and intermolecular zero-order crosslinks in APOA1. EDC reacts with the amino group of lysine resides that are close to the carboxylic acid group of aspartate and glutamate to form an amide bond. Thus, EDC crosslinks identify salt bridges in structures. The crosslinked r-HDLs were reisolated by high-resolution size exclusion chromatography to eliminate HDL particles that were crosslinked to each other. Importantly, all crosslinking reactions were carried out at low concentrations of EDC in phosphate-buffered normal saline at pH 6.5, which more closely mimics physiological conditions than those used to crystallize proteins.

After the crosslinked APOA1 was digested, the resulting peptide mixture was fractionated by capillary liquid chromatography and analyzed by tandem MS/MS. To distinguish between inter- and intramolecular crosslinks of APOA1 in r-HDL, we used a 1:1 mixture of human [^14^N]APOA1 (light, L) and [^15^N]APOA1 (heavy, H) (isotopic purity >99%) to generate the particles.^20,38^ Three combinations of crosslinks are possible: L-L, L-H, and H-H. The L and H forms of APOA1 are chemically identical but differ in molecular mass, making intramolecular and intermolecular crosslinks readily distinguishable in MS1 scans. For intra-protein crosslinks, where the protein is linked to itself, only L-L and H-H forms are detected (relative abundance ∼1:1). For inter-protein crosslinks, LL, L-H, and H-H peptides are detected (relative abundance ∼1:2:1) (**Supplemental Material, Fig. S1 and S2**).^20,38^

### Different sizes of r-HDL exhibit distinct patterns of intramolecular and intermolecular crosslinks between peptides in the N-terminal and C-terminal regions of APOA1

This approach identified 34 intramolecular and 31 intermolecular crosslinks in the four sizes of r-HDL (**Supplemental Material, Table S2**). Similar numbers of intramolecular crosslinks were detected in the three largest particles (r-HDL-120, 8 crosslinks; r-HDL-96, 6 crosslinks; r-HDL-88, 7 crosslinks). In contrast, we identified twice as many intramolecular crosslinks in the r-HDL-80 particle (13 crosslinks). These observations indicate that APOA1 has greater conformational freedom in the smallest r-HDL particles than in the other sizes of HDL.

### Probing the behavior of HDL particles over time with molecular dynamic simulations

To investigate how APOA1 conformation and mobility vary in the different sizes of r-HDL, we used a computational method called molecular dynamics (MD). Here trajectories of systems modeling r-HDL (2 APOA1 bound to a nanodisc composed of POPC and ∼10% cholesterol, and surrounded by water) are generated for multiple microseconds. The result is a series of snapshots of the dynamic evolution for all of the atoms in the system.

We previously used this approach to generate trajectories of r-HDL-100 and r-HDL-120 particles (100 Å and 120 Å diameter particles, Supplemental Material).^27^ To generate the double belt models for r-HDL-80 and r-HDL-90 particles (80 Å and 90 Å diameter particles), we ran MD simulations after removing cholesterol and POPC from the computer generated r-HDL-100 particle (Supplemental Material). We then determined whether the crosslinks we identified in APOA1 in the different sizes of r-HDL were consistent with the double belt model.^34^

To perform this analysis, we compared inter- and intramolecular distances between Cα atoms in the models, using an HDL contact map to plot a detected peptide by the position of its two amino acids in APOA1’s sequence. The maximum distance between the backbone Cα atoms of amino acids in the crosslinked peptides is the sum of the length of two side chains plus the length of the amide bond formed by EDC (10.5 Å for K–D linkage and 12.1 Å for K–E). **Fig. 2** shows the position of each crosslink in the contact maps for the simulated belt structure for each size of HDL. The cutoff radius for the crosslink residing in the double belt model was 15.1 Å (12.1 Å for the K–D crosslink plus a 3 Å motion averaging factor).^27^

**Figure 2.**
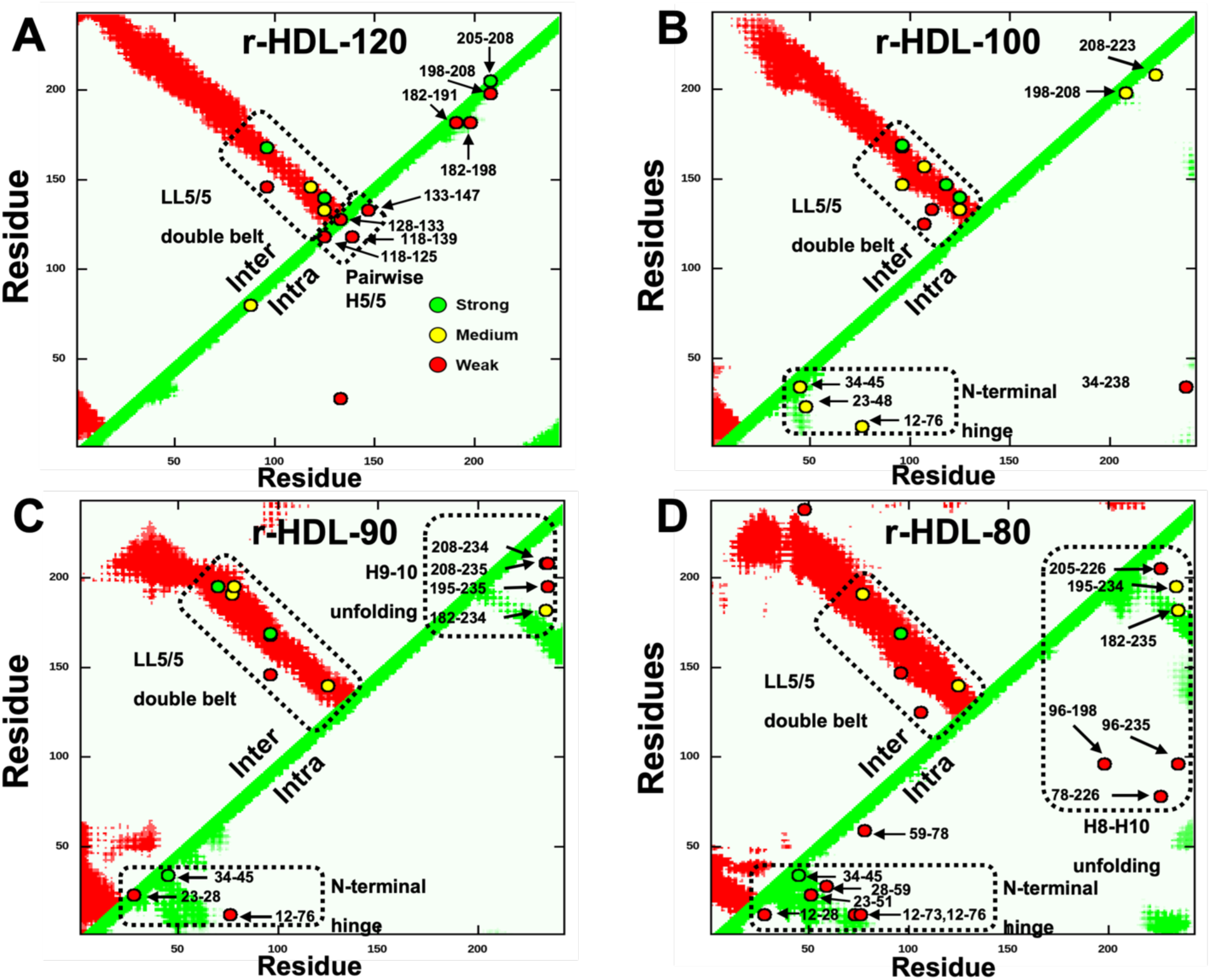
Contact maps of the intermolecular (Inter) and intramolecular (Intra) APOA1 crosslinks detected by MS/MS in different sizes of r-HDL. *Red regions* and *green regions* indicate the allowable distance of intermolecular and intramolecular peptide contacts (15.1 Å), respectively, in a molecular dynamics simulation of the LL5/5 double-belt model of APOA1.^27^ Crosslinks (o) between APOA1 residues are labeled. Semi-quantitative estimates of the strengths of interactions between residues were based on ion currents (**Supplemental Material, Table S2**), and they are indicated by the colors of the circles (green, strong; yellow, medium; red, weak). Note that we detected multiple intramolecular crosslinked peptides in the helix 8 to helix 10 region and helix 9 to helix 10 region of the C-terminus of APOA1 of r-HDL-80 and r-HDL-90 particles, respectively, that are inconsistent with the classic double belt model. This indicates that the C-terminus of APOA1 has increased conformational freedom and does not assume the double belt conformation in that region. In contrast, the intramolecular crosslinked peptides detected in that region of the two largest sizes of HDL are consistent with the double belt model.

Only one of the crosslinks detected in the largest particle (r-HDL-120) was inconsistent with the double belt model of HDL (**Fig. 2A**; intermolecular crosslinks, red regions; intramolecular crosslinks, green regions). Just two of the crosslinks in the r-HDL-100 particle were inconsistent (**Fig. 2B**). In contrast, 7 and 9 crosslinks in the r-HDL-90 and r-HDL-80 particles were inconsistent with the prototypical double belt model. In r-HDL-90, four of the 7 crosslinks were in the C-terminus of APOA1; in r-HDL-80, six of the 9 crosslinks were in the C-terminus (**Fig. 2C-D**). The large number of crosslinks in the C-terminus of the two smallest particles (r-HDL-90 and r-HDL-80) indicates that this region is more loosely organized than in the larger particles and thus underwent a larger number of cross-linking reactions in the experiment.

These observations indicate that the central region of the APOA1 dimer is organized as a double belt in all the sizes of r-HDL we studied. In contrast, the N-terminal and C-terminal regions differ according to particle size. Specifically, the C-termini of APOA1 are markedly more mobile in the two smallest particles. Many of the observed crosslinks in the N-termini of APOA1 of the three smallest HDLs also fell outside the contact zones predicted by the double belt model (**Fig. 2B-D**), which is consistent with the proposal that the N-terminus APOA1 in r-HDL-100 is hinged.^27^

### The C-termini of small HDL particles exhibit greater mobility than those of large HDL particles

Figure 3A-H illustrates the conformational states of APOA1 obtained by MD simulations of the HDL particles of varying sizes, with lipids and water excluded from the image for clarity. In the case of r-HDL-120 particles, the two proteins adopt a predominantly double-belt arrangement, although some displacement of the C-terminal helices (H10A and B) occurs.^27^ Disorder in the N-terminal region (residues 1-43) is noticeable in r-HDL-100 particles, and both the N- and C-termini lose their double-belt characteristics in r-HDL-90 particles. Both C-termini are flipped off the lipid edge in the r-HDL-90 particles (Fig. 3E**,F**).

**Figure 3.**
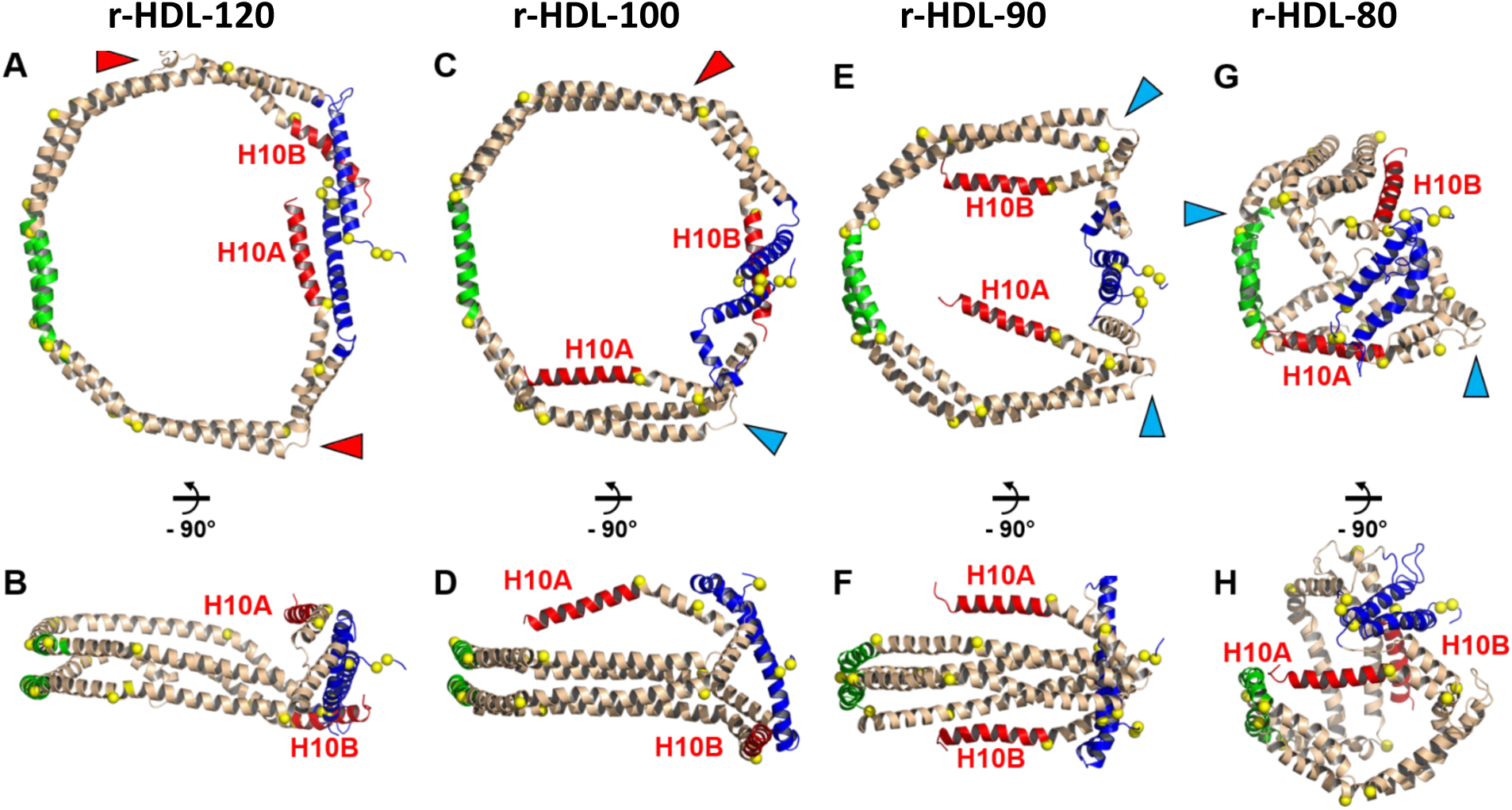
Comparison of stepwise conformational changes in APOA1 between r-HDL-120, r-HDL-100, r-HDL-90 and r-HDL-80 particles. Cartoon representations: **blue,** N-terminal 43 (residues 1-43); **green**, pairwise helix 5 (residues 121-143); **red**, helix 10 (residues 220-243). **Red arrowheads** show the partially unfolded H7-H8 junctions. **Blue arrowheads** show the hairpin coil that allows helix 10 (**H10A** and **H10B**) to fold onto the POPC headgroup surface. **A.** Top view of r-HDL-120. **B.** Side view of r-HDL-120. **C.** Top view of r-HDL-100. **D.** Side view of r-HDL-100. **E.** Top view of r-HDL-90. **F.** Side view of r-HDL-90. **G.** Top view of r-HDL-80. **H.** Side view of r-HDL-80. Note that APOA1 in the two largest HDL particles (**A-D**; r-HDL-120 and -100) has a conformation that is strongly lipid-associated and consistent with the classic double belt model. In contrast, this structure is absent in both C-termini (**H10A,B**) of APOA1 in the two smallest HDL particles (**E-H**; r-HDL-90 and -80).

The structure of the smallest particle, r-HDL-80, markedly differs from that of the larger r-HDL particles. The smallest particles are shaped like a spherical micelle, deviating from the disk-like structure, and the double-belt arrangement is largely absent except for helix 5 (green) (Fig. 3G**,H**). There is also significant unwinding and displacement of the C-terminal helices.

It is important to note that the images (Fig. 3) represent snapshots of individual simulations at specific time points and do not capture the complete flexibility of the terminal helices. Although the simulations of the large r-HDL-100 and r-HDL-120 particles show that the C-termini remain associated with the lipid surface, they could detach in the presence of a protein such as ABCA1. Thus, these images represent low energy states but not the sole states achievable by these systems.^28^ It is also worth noting that the time scale, potential energy functions, and restraints employed in the all-atom and coarse-grained simulations tend to maintain the helical structure of the protein residues.

### Clinical characteristics of LCAT-deficient and control subjects

Like the reconstituted HDL particles used in our model system studies, the extra-small and small HDL particles in LCAT-deficient subjects are discoidal and composed largely of APOA1, free cholesterol, and phospholipid.^39,40^ We therefore used serum HDL and HDLs isolated from control and LCAT-deficient subjects to investigate the relevance of our model system studies to human HDL.

We studied three groups of subjects: 14 controls, 6 subjects with heterozygous LCAT deficiency (LCAT+/-), and 6 with LCAT deficiency (LCAT-/-).^31^ Two of the LCAT-/- subjects were unrelated to the subjects in the family study. The three groups had similar ages, percentages of females, and plasma LDL-C levels (**Table 1**). Compared to the LCAT+/+ subjects, the LCAT+/- subjects had significantly lower plasma HDL-C levels, as did the LCAT-/- subjects (*P*=0.0005). Plasma triglyceride levels were not significantly different between the groups (*P*=0.28; Mixed effect model and Tukey-Kramer post-tests). LCAT activity was undetectable in the LCAT-/- subjects (0 nmol/mL per h), with significantly higher levels in the LCAT+/- subjects and LCAT+/+ subjects (*P*<0.0001).

**Table 1.** Clinical characteristics of the subjects in the family study.

| Characteristic | Controls<br>(LCAT+/+)<br>n=14 | Heterozygotes<br>(LCAT+/-)<br>n=6 | Homozygotes<br>(LCAT-/-)<br>n=4 | P-value |
| --- | --- | --- | --- | --- |
| Age (years) | 49±20 | 54±16 | 47±18 | 0.81 |
| Female/Male (n) | 5/9 | 2/4 | 4/2 | --- |
| HDL-C (mg/dl) | 71±9 | 40±11 | 9.2±6.1 | .0005 |
| LDL-C (mg/dl) | 114±32 | 109±34 | 111±74 | 0.22 |
| Triglycerides (mg/dl) | 111±61 | 121±30 | 253±130 | 0.28 |
| LCAT activity<br>(nmol/mL per h) | 42±9 | 20±13 | 0±0 | <.0001 |

*P*-values are from a mixed effect model and Tukey-Kramer post-tests.

### Extra-small HDL (XS-HDL) and small HDL (S-HDL) particles are enriched in LCAT-deficient subjects

We used calibrated IMA to quantify total HDL (Total HDL) and four sizes of HDL particles: extra-small HDL (XS-HDL), small HDL (S-HDL), medium HDL (M-HDL), and large HDL (L-HDL) (Fig. 4A**; Supplemental Material, Table S3**). This method for quantifying HDL-P yields a stoichiometry of APOA1 and sizes and relative abundances of HDL subspecies that agree well with those determined by non-denaturing gradient gel electrophoresis and analytical ultracentrifugation.^23,41^ In the LCAT+/+ subjects, M-HDL (mean diameter, 9.2±0.1 nm) was the most abundant particle population; it accounted for ∼50% of total HDL (Fig. 4A). All four sizes of HDL were detected in all the control subjects. In contrast, subjects with partial LCAT deficiency had elevated levels of XS-HDL (mean diameter, 7.8±0.1 nm) and S-HDL (mean diameter, 8.4±0.1 nm diameter) particles (Fig. 4A). Subjects with complete LCAT deficiency exhibited only XS-HDL (Fig. 4A).

**Figure 4.**
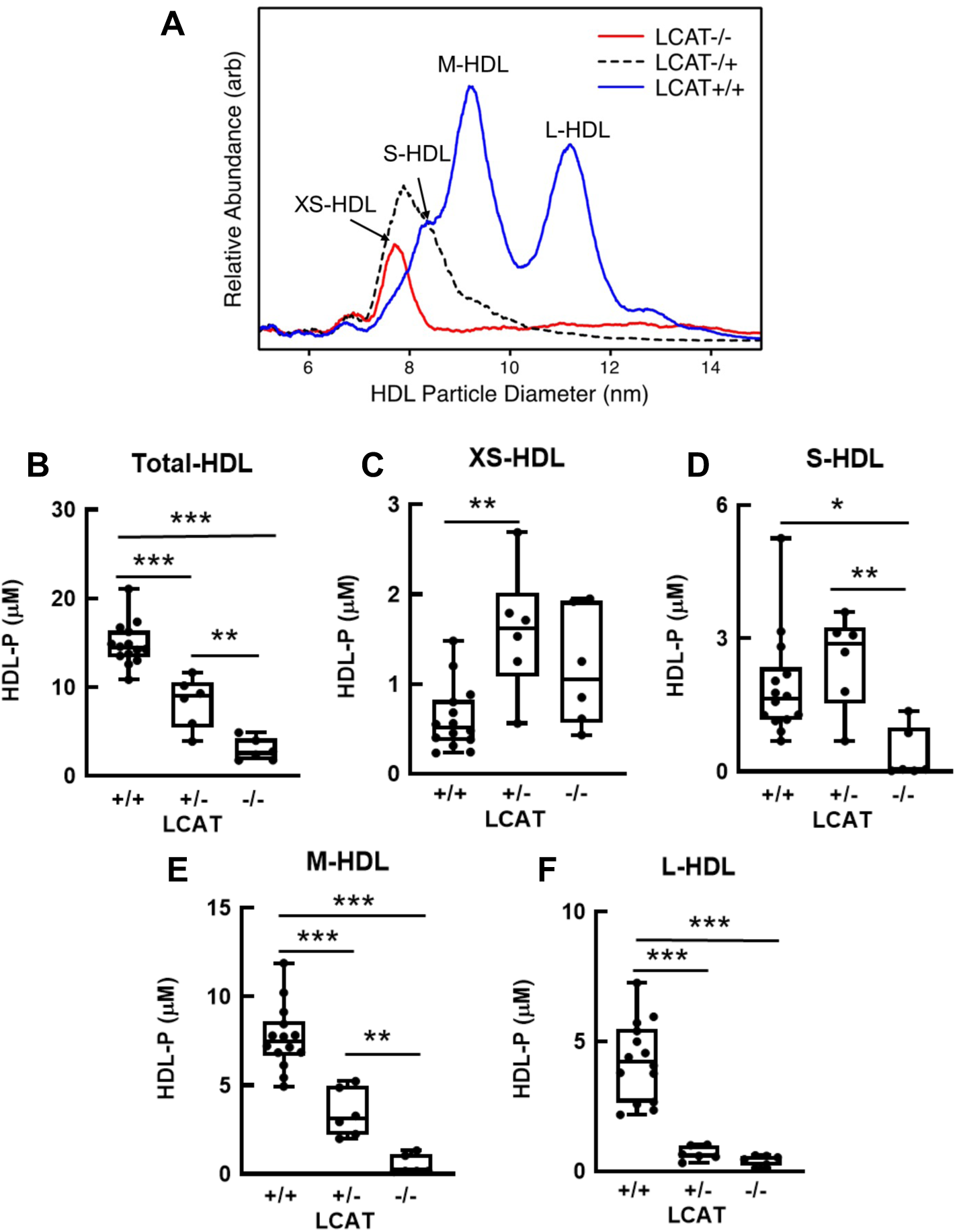
Quantification of total HDL and HDL subspecies in LCAT-deficient (-/-), LCAT-heterozygous (+/-), and control (+/+) subjects. (**A**) Representative size and concentration profiles of HDL isolated from LCAT-deficient (-/-), LCAT-heterozygous (+/-), and control (LCAT +/+) subjects. (**B-F**) HDL isolated by ultracentrifugation from plasma (d=1.063-1.21 g/mL) was analyzed by calibrated IMA. The mean HDL subspecies sizes were: extra-small HDL (XS-HDL) 7.8 nm; small HDL (S-HDL) 8.4 nm; medium HDL (M-HDL) 9.2 nm; large HDL (L-HDL) 10.9 nm. Arb, arbitrary units. HDL isolated from plasma by ultracentrifugation was subjected to calibrated IMA. The sizes (mean, standard deviation) of the HDL subspecies were: extra-small HDL, 7.8±0.1 nm; small HDL, 8.4±0.1 nm; medium HDL, 9.2±0.1 nm; large HDL, 10.9±0.2 nm. The number of subjects was: LCAT+/+, n=14; LCAT+/-, n=6; LCAT-/-, n=6. *P*-value, one-way ANOVA with Tukey-Kramer post-tests. *** *P*<0.001, ** *P*<0.01, * *P*<0.05.

The total concentration of HDL particles and the distribution of the different sizes of HDL also differed significantly among the three groups (Fig. 4B**-F****; Supplemental Material, Table S3**). Mean Total-HDL-P levels in control subjects were 5.1 times higher than those in LCAT-/- subjects and 1.8 times higher than those in LCAT+/- subjects (*P*<0.0001). This reflected significantly lower levels of both M-HDL and L-HDL in subjects with complete or partial LCAT deficiency (*P*<0.0001 for both M-HDL and L-HDL). In contrast, mean levels of XS-HDL were higher in both LCAT-/- and LCAT+/- subjects than in control subjects. LCAT-/- and LCAT+/- subjects had similar levels of XS-HDL and L-HDL.

### Serum HDL from LCAT-deficient subjects has normal macrophage and ABCA1 cholesterol efflux capacity despite low total HDL-P

We used serum HDL (APOB-depleted serum) to quantify the subjects’ CEC, as described by Rothblat et al.^10,11^ Macrophage and ABCA1 CEC were evaluated using J774 macrophages stimulated with cAMP or BHK cells with mifepristone-inducible expression of human ABCA1. CEC, quantified as the difference in cholesterol efflux with and without induction of ABCA1, was a linear function of serum HDL concentration and incubation time. A mixed effect model demonstrated that macrophage CEC and ABCA1 CEC did not differ significantly between LCAT-/- and control subjects (*P*>0.3).

To begin to identify the HDL subpopulations that drive macrophage CEC and ABCA1 CEC, we correlated the CEC of serum HDL with the particle concentration of each HDL subpopulation from all the study subjects (**Supplemental Fig. S3**). Macrophage CEC only correlated positively and strongly with the concentration of XS-HDL particle concentration (r=0.55, *P*=0.004). These observations suggest that XS-HDL is an important driver of cellular cholesterol export from both macrophages and through the ABCA1 pathway.

### Small and extra-small HDL particles are the major promoters of macrophage CEC and ABCA1 CEC in both LCAT-deficient subjects and control subjects

To further investigate how XS-HDL promotes CEC, we used ultracentrifugation and high-resolution size exclusion chromatography to isolate XS-HDL from LCAT-/- subjects. We then compared the CEC activity of XS-HDL with that of four sizes of HDL isolated from LCAT +/+ subjects. The mean diameters of the isolated HDLs were 8.4 nm, 8.8 nm, 9.2 nm, and 14 nm (Fig. 5A; size distributions), respectively.

**Figure 5.**
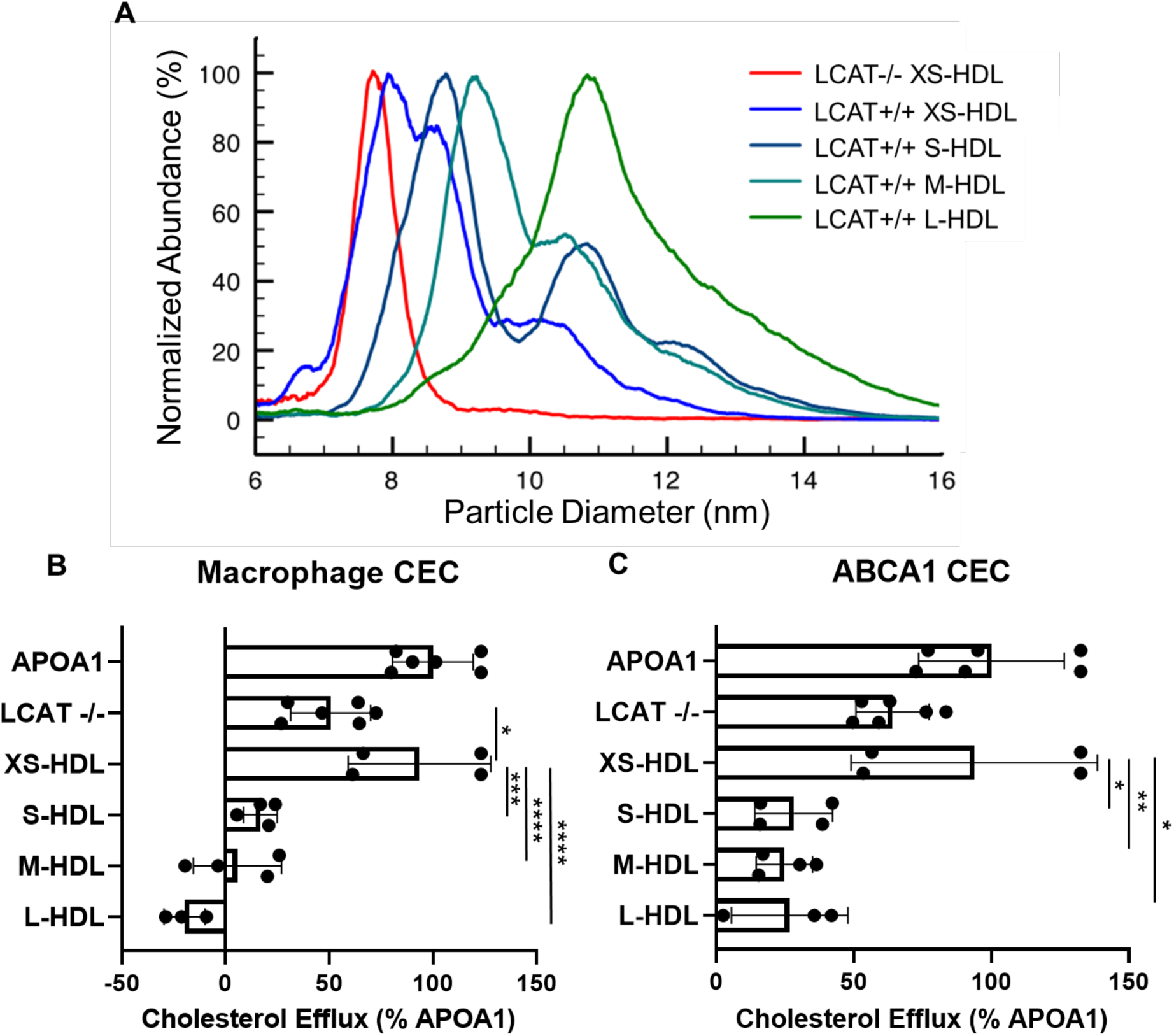
Calibrated IMA (A) and CEC (B) of HDL isolated from plasma of LCAT-deficient (LCAT-/-) and control (XS-HDL, S-HDL, M-HDL, L-HDL) subjects. (**A)** Representative IMA size profiles of isolated HDL. To facilitate comparison of the particles’ size distributions, the height of each isolated HDL fraction was set to 100%. The diameters of the isolated HDLs of LCAT-/- subjects and control subjects were: LCAT-/-, 7.8±0.1 nm; XS-HDL, 8.1±0.2 nm; S-HDL, 8.8±0.1 nm; M-HDL, 9.8±0.2 nm; L-HDL, 11.1±0.2 nm. Note that isolated XS-HDL is composed of both XS-HDL and S-HDL particles. (**B, C**) ABCA1-mediated cholesterol efflux capacity (CEC) of HDL isolated from LCAT-/- subjects and control subjects. Macrophage CEC and ABCA1 CEC of were quantified with [^3^H]cholesterol-labeled J774 macrophages and baby hamster kidney cells after a 4-h incubation. Expression of ABCA1 was induced with cAMP and mifepristone, respectively. Cholesterol efflux was calculated as the percentage of radiolabel in the medium of the cells divided by the total radioactivity of the medium and cells. CEC was quantified as the difference in cholesterol efflux of cells with and without induced expression of ABCA1. Isolated HDLs were included in the media of the cells at equal particle concentrations. CEC of HDLs was normalized to CEC of cells exposed to 10 µg/mL of APOA1. *P*-value: one-way ANOVA with Tukey-Kramer post-tests. \*\*\**P*<0.001, ** *P*<0.01, * *P*<0.05.

We quantified macrophage and ABCA1 CEC as described above for serum HDL. Importantly, we incubated the cells with equimolar concentrations of isolated particles of each size of HDL. CEC was a linear function of HDL particle concentration and the incubation time used in the assays. XS-HDL isolated from LCAT-/- subjects, composed almost exclusively of 7.8 nm diameter particles (XS-HDL-sized), strongly promoted both macrophage and ABCA1 CEC. However, it was less potent than 8.4 nm XS-HDL particles isolated from LCAT +/+ subjects (Fig. 5B), which were composed of approximately equimolar amounts of XS-HDL and S-HDL (as determined by calibrated IMA). On a molar basis, 8.4 nm HDL isolated from control subjects was as effective as lipid-free APOA1 in promoting both macrophage CEC and ABCA1 CEC. On a molar basis, XS-HDL isolated from LCAT-/- subjects promoted macrophage CEC much more effectively than 8.8 nm HDL, 9.2 nm HDL, or 14 nm HDL isolated from control subjects, and the differences were significant (*P*=0.0002 for 8.8 nm HDL, *P*<0.0001 for 9.2 nm HDL, and *P*<0.0001 for 14 nm HDL). We obtained similar results when we determined how effectively the different sizes of isolated HDL promoted ABCA1 CEC (Fig. 5C).

### LCAT converts small HDLs into large HDLs, markedly reducing CEC

To test the hypothesis that LCAT is one important factor controlling the CEC of circulating HDL, we incubated control plasma and LCAT-deficient plasma with or without recombinant human LCAT at 37°C for 2 h, stopped the LCAT reaction with DTNB, and quantified ABCA1 CEC and HDL-P, using calibrated IMA. Control experiments demonstrated that DTNB had no impact on quantification of ABCA1 CEC.

Before incubation with LCAT, the major HDL species in the LCAT-/- subjects was XS-HDL. In contrast, all 4 sizes of HDL were observed in the control subjects; M-HDL was the most abundant species. LCAT treatment decreased the ABCA1 CEC of both control plasma and LCAT-/- plasma by ∼50% (Fig. 6A). LCAT converted virtually all XS-HDL and most S-HDL particles into larger HDLs in plasma of both control and LCAT-deficient subjects (Fig. 6B). In plasma treated with LCAT, free cholesterol markedly decreased while total cholesterol did not change significantly (**Fig 6C-D**).

**Figure 6.**
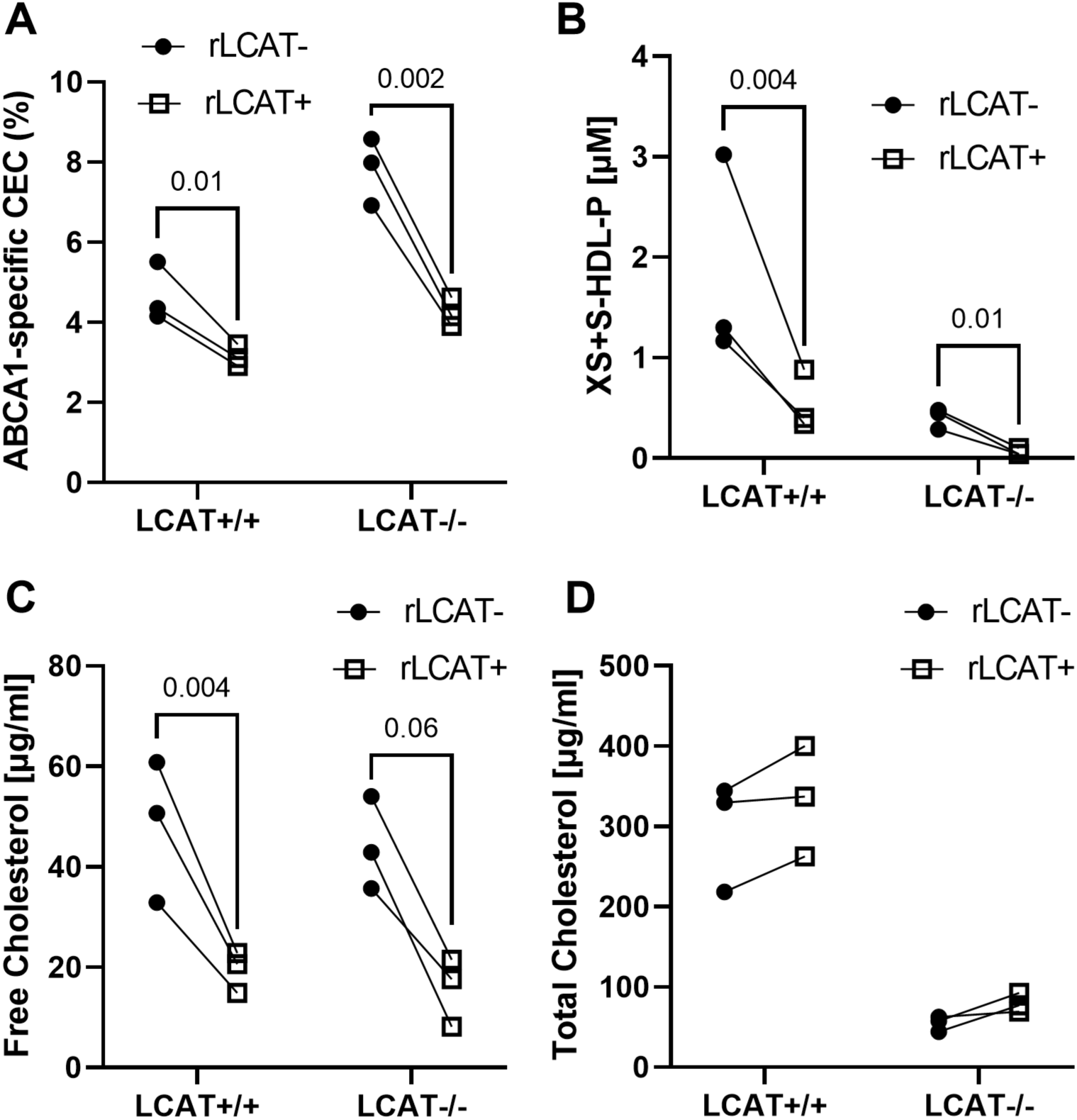
ABCA1 CEC (A), HDL particle size distribution (B), free cholesterol (C) and total cholesterol (D) content of control and LCAT-deficient plasma incubated with LCAT. Control plasma (N=3) and LCAT-deficient plasma (N=3) were incubated with and without recombinant human LCAT (**rLCAT+** and **rLCAT-**; 50 μg/mL) for 1 h at 37^⁰^C. The LCAT reaction was stopped with 2 mM DTNB and cooling on ice. Control studies demonstrated that DTNB did not alter the CEC of plasma. DTNB was omitted from plasma used to quantify cholesterol levels because it interfered with the enzymatic assay. ABCA1 CEC of plasma was quantified using [^3^H]cholesterol-labeled BHK cells as described in the legend to Fig. 5. *P*-values, ratio-t test.

Collectively, these observations support the proposal that small HDLs are the major HDL species promoting both ABCA1 and macrophage CEC.

## Discussion

To investigate the mechanisms that regulate HDL’s ability to promote cholesterol efflux by the ABCA1 pathway, we quantified the CEC of four different sizes of r-HDLs. As with human HDL,^7,8^ the smallest r-HDL particles were the strongest promoters of cholesterol efflux. Chemical crosslinking followed by MS/MS analysis showed that twice as many intramolecular crosslinked peptides had formed in the smallest r-HDL than in the three larger sizes, indicating that APOA1 had markedly higher mobility. When we plotted the positions of the chemically crosslinked peptides on an HDL contact map, virtually all the peptides detected in the two largest r-HDL particles were consistent with molecular dynamics simulations of the double belt model of APOA1. In this model, the helical repeats of two APOA1 molecules assume an anti-parallel helical structure that forms a bundle surrounding the edges of discoidal HDL. Because the helical bundle is amphipathic and has high lipid affinity,^42^ these observations strongly suggest that most of the APOA1 in the two largest HDL particles is bound to lipid and therefore would not be accessible to ABCA1.

The two smallest HDL particles showed a different pattern of chemically crosslinked peptides: the peptides of APOA1’s central region were consistent with the double belt model, but those at the C-terminus were not (i.e., the C-termini in the APOA1 dimer are not in a helical bundle). Moreover, the smallest HDL had the largest number of detectable chemically crosslinked peptides in the C-terminus of APOA1. Taken together, these data indicate that the C-terminus of APOA1 in the small HDL particles have enhanced conformational mobility, likely due to a loss of overall helicity. Because lipid interaction is a major driver of helical formation in APOA1, it is conceivable that the C-terminus is detached from the lipid surface in the two smallest r-HDLs as observed in the MD simulations of those particles.

The C-terminus of APOA1 plays a critical role in promoting cholesterol export by ABCA1.^42–44^ We envision two factors that drive its increased mobility and loss of helical structure in small HDLs. First, crowding, as small HDL’s limited surface area might not accommodate all of APOA1’s helices, preventing the C-termini of APOA1 from lying on the particle’s surface. Second, extreme surface curvature, which could prevent APOA1’s C-terminal domain from fully interacting with lipid because it cannot turn sharply enough to lie down on the surface. In this model, the two antiparallel C-termini are “flipped” off the surface of smaller HDLs (Fig. 7), where their increased mobility and freedom from lipid binding promote their engagement with ABCA1. In contrast, the C-termini of larger HDLs are strongly bound to lipid and unable to interact productively with ABCA1. On a molar basis, the smallest r-HDL and human HDL particles were as efficient at promoting cholesterol efflux by the ABCA1 pathway as was lipid-free APOA1, which is consistent with this model.

**Figure 7.**
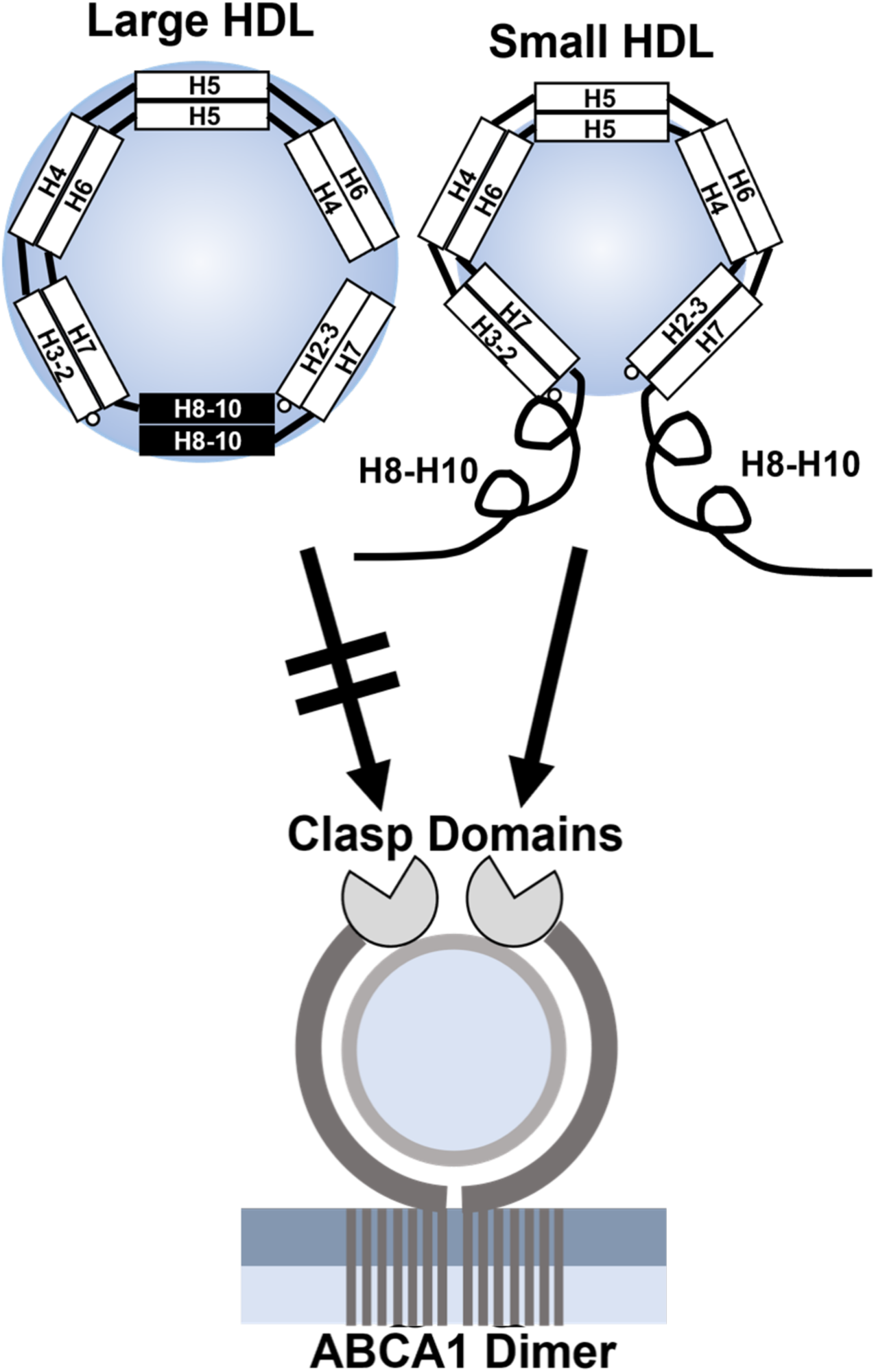
The “flipped ends” model for the increased ABCA1 activity of small HDLs. In large HDL particles, the C-termini of the APOA1 dimer are in antiparallel helical bundles that are amphipathic and strongly associated with lipid. In small HDL particles, the reduced surface area and high surface curvature force the C-termini off the particles, increasing their mobility. The termini also are less lipid-associated because APOA1 loses its amphipathic double belt structure. Decreased lipid association and increased mobility of the C-termini (helices H8–10) promote the engagement of APOA1 with the clasp domains of ABCA1, stimulating cholesterol export from the cell. An alternative hypothesis is that the C-termini of APOA1 promote microsolubilization of phospholipids and cholesterol from phospholipid-rich domains in the plasma membrane of cells (see Discussion).

We suggest that the smaller, less lipidated HDL particles may not be fully “filled” with lipid and remain effective substrates for ABCA1. At some point between diameters of 80 and 90 Å, the APOA1 C-terminus finds room to associate with the surface of the particle, shutting down additional lipid accumulation by ABCA1 and possibly promoting the release of a more ‘mature’ particle. In this scheme, the APOA1 C-terminus functions as a lipid level switch that defines the maximal size of HDL particles formed by ABCA1.

An alternative hypothesis for the role of the C-terminus of APOA1 in promoting CEC is that it involves binding of the protein to phospholipid-rich domains in the plasma membrane of cells,^45,46^ which in turn promotes phospholipid and cholesterol efflux. Consistent with this, deletion of helices H8-H10 decreases binding to lipid vesicles but has little impact on cross-linking of radiolabeled APOA1 to ABCA1 on cells. However, our recent studies^47^ suggest that the lipid-affinity of APOA1 plays a role in promoting the movement of phospholipids from the outer-leaflet of the plasma membrane into a hydrophobic tunnel in the interior of the extracellular domain of ABCA1.

To test the relevance of our model system studies to human HDL, we used serum HDL and HDL isolated from LCAT-deficient carriers. Like r-HDLs, the latter is discoidal and composed of APOA1, free cholesterol, and phospholipid. We found that total HDL particle concentration was markedly lower in subjects who completely lacked LCAT activity. The major HDL subspecies in those subjects was XS-HDL, which is very similar in size (78 Å in diameter) to the smallest r-HDL-80 particles used in our model system studies (80 Å). In subjects who were only partially LCAT-deficient, the major HDL subspecies were XS-HDL and S-HDL. In subjects with normal LCAT activity, we detected all four sizes of HDL; M-HDL was the major species. Even though serum from the LCAT-deficient subjects had very low HDL particle concentrations, ABCA1 CEC and macrophage CEC were similar to that of the controls, strongly suggesting that the ABCA1-specific activities of extra-small and small HDL were greater than those of the larger HDL particles. Consistent with this proposal, macrophage and ABCA1 CEC strongly and positively correlated with the concentration of XS-HDL in plasma.

To confirm these ideas, we used isolated HDL from subjects with and without complete LCAT deficiency. Then we determined macrophage and ABCA1 CEC activity at equal particle concentrations. The isolated particles had diameters of 8.4 nm, 8.8 nm, 9.2 nm, and 14 nm—very similar to the sizes of XS-HDL, S-HDL, M-HDL, and L-HDL in plasma as quantified by calibrated IMA. It is important to note that the isolated HDLs did not precisely mimic the size distributions of the HDL subclasses in plasma from the LCAT-deficient and control subjects. Extra-small HDL isolated from control subjects, which was a mixture of XS-HDL and S-HDL, had the highest macrophage and ABCA1 CEC specific activities; they were about 4–5-fold greater than those of the three larger sizes of isolated HDL. XS-HDL isolated from the LCAT-deficient subjects did not contain S-HDL and was less active than XS-HDL isolated from the control subjects; macrophage CEC and ABCA1 CEC of the isolated HDLs were ∼3-fold greater than for the larger sizes of HDL. Lipid-free APOA1 was not detectable in the HDL used for these studies because the particles were isolated by both ultracentrifugation and high-resolution size exclusion chromatography.

These data suggest that both XS-HDL and S-HDL are the major contributors to macrophage and ABCA1 CEC. This hypothesis is strongly supported by the demonstration that incubating control plasma and LCAT-deficient plasma with LCAT converted small HDLs into large HDLs and markedly diminished ABCA1 CEC. These observations are remarkably concordant with animal studies, which demonstrated that ABCA1 CEC of plasma HDL was increased in LCAT-/- and LCAT +/- mice.^16^ Moreover, over-expression of LCAT significantly reduced macrophage cholesterol efflux by plasma. Taken together, these observations suggest that XS-HDL and S-HDL, which typically represent 20%-30% of total HDL, are key mediators of ABCA1 CEC and perhaps cardioprotection.

CSL-112, a reconstituted HDL particle that promotes the formation of small and lipid-poor APOA1 particles,^18,19^ is being tested in a large, randomized study to determine if it reduces the risk of CVD events in post-MI patients. The demonstration that CLS-112 lowers incident CVD would strongly support the proposal that small HDLs are critical in cardioprotection in humans.

Our demonstration that small and extra-small HDL particles potently promote cholesterol efflux from macrophages raises the possibility that increased LCAT activity, which converts smaller HDL particles into larger, cholesteryl ester-rich particles, is a risk factor for atherosclerosis.^48^ Consistent with this suggestion, overexpression of LCAT in mice failed to increase reverse cholesterol transport from macrophages to bile.^16^ Serum HDL from mice that overexpressed LCAT were less able to promote cholesterol efflux from macrophages by the ABCA1 pathway than control mice.^16^ However, studies of the relationships of LCAT to CVD risk in humans have yielded mixed results.^49,50^

One limitation of our investigations is the small number of subjects in our study of LCAT deficiency. However, the large differences in the concentrations of the various sizes of HDL in the different groups of subjects and the consistency of the results with serum HDL and isolated HDL strongly support the proposal that XS-HDL and S-HDL promote cholesterol efflux from macrophages by the ABCA1 pathway. Another limitation is that the size distributions of the isolated HDLs overlapped to some degree, reflecting the limited resolution of size-exclusion chromatography. Nonetheless, the mean sizes of the isolated HDLs were well separated, and the particle distributions were clearly distinct from one another.

In summary, both our experiments and molecular dynamics simulations support the proposal that small HDL particles are potent ligands for promoting cholesterol efflux from macrophages by the ABCA1 pathway. In future studies, it will clearly be important to determine whether XS-HDL and S-HDL predict CVD risk in humans, if LCAT mass and/or activity associate with HDL size, and if risk prediction is independent of HDL-C.

## Acknowledgments

We thank Dr. Priska von Haller and the Proteomics Resource (UWPR95794, University of Washington) for technical support and Dr. Fabrizio Veglia for statistical advice. Molecular dynamics simulations were performed at the National Institutes of Health (NIH), Bethesda, MD (BIOWULF and Lobos clusters) and on the Anton2 supercomputer. Access to Anton 2 was generously provided by D.E. Shaw Research.

## Data availability

The authors confirm that the data supporting the findings of this study are available within the article and/or its supplementary materials.

## Sources of Funding

This work was supported by awards from the NIH: T32HL007828, R01HL149685, R35HL150754, P01HL151328, P01HL128203, P30DK017047, R01HL153118, R01HL155601, R01HL144558, and R01HL149685; the American Heart Association (15POST22700033); and the Intramural Program of the National Heart Lung and Blood Institute of the NIH. Anton 2 time was provided by the Pittsburgh Supercomputing Center (NIH R01GM116961).

## Disclosures

K.E.B. serves on the Scientific Advisory Board of Esperion Therapeutics.

## Notes

### Author Declarations

Ethics Committees/IRBs of Milano Area C, Italy (approval number 446-092014), New York University (approval number I14-01537) and the University of Washington (approval number STUDY00012123), gave ethical approval for this work. Ethics Committees in Italy are set up in structures identified by regions rather than by institutions.

## References

1. Gordon DJ and Rifkind BM. High-density lipoprotein--the clinical implications of recent studies. N Engl J Med. 1989;321:1311–6.

2. Rader DJ and Hovingh GK. HDL and cardiovascular disease. Lancet. 2014;384:618–625.

3. Rader DJ and Tall AR. The not-so-simple HDL story: Is it time to revise the HDL cholesterol hypothesis? Nat Med. 2012;18:1344–6.

4. Heinecke J. HDL and cardiovascular-disease risk--time for a new approach? N Engl J Med. 2011;364:170–1.

5. Rader DJ, Alexander ET, Weibel GL, Billheimer J and Rothblat GH. The role of reverse cholesterol transport in animals and humans and relationship to atherosclerosis. J Lipid Res. 2009;50 Suppl:S189–94.

6. Oram JF and Heinecke JW. ATP-binding cassette transporter A1: a cell cholesterol exporter that protects against cardiovascular disease. Physiol Rev. 2005;85:1343–72.

7. Du XM, Kim MJ, Hou L, Le Goff W, Chapman MJ, Van Eck M, Curtiss LK, Burnett JR, Cartland SP, Quinn CM, Kockx M, Kontush A, Rye KA, Kritharides L and Jessup W. HDL particle size is a critical determinant of ABCA1-mediated macrophage cellular cholesterol export. Circ Res. 2015;116:1133–42.

8. He Y, Ronsein GE, Tang C, Jarvik GP, Davidson WS, Kothari V, Song HD, Segrest JP, Bornfeldt KE and Heinecke JW. Diabetes Impairs Cellular Cholesterol Efflux From ABCA1 to Small HDL Particles. Circ Res. 2020;127:1198–1210.

9. Yvan-Charvet L, Wang N and Tall AR. Role of HDL, ABCA1, and ABCG1 transporters in cholesterol efflux and immune responses. Arteriosclerosis, thrombosis, and vascular biology. 2010;30:139–43.

10. de la Llera-Moya M, Drazul-Schrader D, Asztalos BF, Cuchel M, Rader DJ and Rothblat GH. The ability to promote efflux via ABCA1 determines the capacity of serum specimens with similar high-density lipoprotein cholesterol to remove cholesterol from macrophages. Arterioscler Thromb Vasc Biol. 2010;30:796–801.

11. Khera AV, Cuchel M, de la Llera-Moya M, Rodrigues A, Burke MF, Jafri K, French BC, Phillips JA, Mucksavage ML, Wilensky RL, Mohler ER, Rothblat GH and Rader DJ. Cholesterol efflux capacity, high-density lipoprotein function, and atherosclerosis. N Engl J Med. 2011;364:127–35.

12. Rohatgi A, Khera A, Berry JD, Givens EG, Ayers CR, Wedin KE, Neeland IJ, Yuhanna IS, Rader DR, de Lemos JA and Shaul PW. HDL cholesterol efflux capacity and incident cardiovascular events. N Engl J Med. 2014;371:2383–93.

13. Saleheen D, Scott R, Javad S, Zhao W, Rodrigues A, Picataggi A, Lukmanova D, Mucksavage ML, Luben R, Billheimer J, Kastelein JJ, Boekholdt SM, Khaw KT, Wareham N and Rader DJ. Association of HDL cholesterol efflux capacity with incident coronary heart disease events: a prospective case-control study. Lancet Diabetes Endocrinol. 2015;3:507–13.

14. Kunnen S and Van Eck M. Lecithin:cholesterol acyltransferase: old friend or foe in atherosclerosis? Journal of Lipid Research. 2012;53:1783–1799.

15. Berard AM, Foger B, Remaley A, Shamburek R, Vaisman BL, Talley G, Paigen B, Hoyt RF, Jr., Marcovina S, Brewer HB, Jr. and Santamarina-Fojo S. High plasma HDL concentrations associated with enhanced atherosclerosis in transgenic mice overexpressing lecithin-cholesteryl acyltransferase. Nat Med. 1997;3:744–9.

16. Tanigawa H, Billheimer JT, Tohyama J, Fuki IV, Ng DS, Rothblat GH and Rader DJ. Lecithin: cholesterol acyltransferase expression has minimal effects on macrophage reverse cholesterol transport in vivo. Circulation. 2009;120:160–9.

17. Calabresi L, Baldassarre D, Castelnuovo S, Conca P, Bocchi L, Candini C, Frigerio B, Amato M, Sirtori CR, Alessandrini P, Arca M, Boscutti G, Cattin L, Gesualdo L, Sampietro T, Vaudo G, Veglia F, Calandra S and Franceschini G. Functional lecithin: cholesterol acyltransferase is not required for efficient atheroprotection in humans. Circulation. 2009;120:628–35.

18. Kingwell BA, Nicholls SJ, Velkoska E, Didichenko SA, Duffy D, Korjian S and Gibson CM. Antiatherosclerotic Effects of CSL112 Mediated by Enhanced Cholesterol Efflux Capacity. J Am Heart Assoc. 2022;11:e024754.

19. Didichenko SA, Navdaev AV, Cukier AM, Gille A, Schuetz P, Spycher MO, Therond P, Chapman MJ, Kontush A and Wright SD. Enhanced HDL Functionality in Small HDL Species Produced Upon Remodeling of HDL by Reconstituted HDL, CSL112: Effects on Cholesterol Efflux, Anti-Inflammatory and Antioxidative Activity. Circ Res. 2016;119:751–63.

20. He Y, Song HD, Anantharamaiah GM, Palgunachari MN, Bornfeldt KE, Segrest JP and Heinecke JW. Apolipoprotein A1 Forms 5/5 and 5/4 Antiparallel Dimers in Human High-density Lipoprotein. Mol Cell Proteomics. 2019;18:854–864.

21. Tubb MR, Smith LE and Davidson WS. Purification of recombinant apolipoproteins A-I and A-IV and efficient affinity tag cleavage by tobacco etch virus protease. J Lipid Res. 2009;50:1497–504.

22. Cavigiolio G, Shao B, Geier EG, Ren G, Heinecke JW and Oda MN. The interplay between size, morphology, stability, and functionality of high-density lipoprotein subclasses. Biochemistry. 2008;47:4770–9.

23. Hutchins PM, Ronsein GE, Monette JS, Pamir N, Wimberger J, He Y, Anantharamaiah GM, Kim DS, Ranchalis JE, Jarvik GP, Vaisar T and Heinecke JW. Quantification of HDL particle concentration by calibrated ion mobility analysis. Clin Chem. 2014;60:1393–401.

24. Caulfield MP, Li S, Lee G, Blanche PJ, Salameh WA, Benner WH, Reitz RE and Krauss RM. Direct determination of lipoprotein particle sizes and concentrations by ion mobility analysis. Clin Chem. 2008;54:1307–16.

25. Guha S, Li M, Tarlov MJ and Zachariah MR. Electrospray-differential mobility analysis of bionanoparticles. Trends Biotechnol. 2012;30:291–300.

26. Rinner O, Seebacher J, Walzthoeni T, Mueller LN, Beck M, Schmidt A, Mueller M and Aebersold R. Identification of cross-linked peptides from large sequence databases. Nat Methods. 2008;5:315–8.

27. Pourmousa M, Song HD, He Y, Heinecke JW, Segrest JP and Pastor RW. Tertiary structure of apolipoprotein A-I in nascent high-density lipoproteins. Proc Natl Acad Sci U S A. 2018;115:5163–5168.

28. Pan AC, Weinreich TM, Piana S and Shaw DE. Demonstrating an Order-of-Magnitude Sampling Enhancement in Molecular Dynamics Simulations of Complex Protein Systems. Journal of Chemical Theory and Computation. 2016;12:1360–1367.

29. Jo S, Kim T, Iyer VG and Im W. CHARMM-GUI: a web-based graphical user interface for CHARMM. J Comput Chem. 2008;29:1859–65.

30. Jones MK, Gu F, Catte A, Li L and Segrest JP. “Sticky” and “promiscuous”, the yin and yang of apolipoprotein A-I termini in discoidal high-density lipoproteins: a combined computational-experimental approach. Biochemistry. 2011;50:2249–63.

31. Calabresi L, Pisciotta L, Costantin A, Frigerio I, Eberini I, Alessandrini P, Arca M, Bon GB, Boscutti G, Busnach G, Frasca G, Gesualdo L, Gigante M, Lupattelli G, Montali A, Pizzolitto S, Rabbone I, Rolleri M, Ruotolo G, Sampietro T, Sessa A, Vaudo G, Cantafora A, Veglia F, Calandra S, Bertolini S and Franceschini G. The molecular basis of lecithin:cholesterol acyltransferase deficiency syndromes: a comprehensive study of molecular and biochemical findings in 13 unrelated Italian families. Arterioscler Thromb Vasc Biol. 2005;25:1972–8.

32. George RT, Abuhatzira L, Stoughton SM, Karathanasis SK, She D, Jin C, Buss N, Bakker-Arkema R, Ongstad EL, Koren M and Hirshberg B. MEDI6012: Recombinant Human Lecithin Cholesterol Acyltransferase, High-Density Lipoprotein, and Low-Density Lipoprotein Receptor-Mediated Reverse Cholesterol Transport. J Am Heart Assoc. 2021;10:e014572.

33. Mendez AJ, Oram JF and Bierman EL. Protein kinase C as a mediator of high density lipoprotein receptor-dependent efflux of intracellular cholesterol. J Biol Chem. 1991;266:10104–11.

34. Segrest JP, Jones MK, Klon AE, Sheldahl CJ, Hellinger M, De Loof H and Harvey SC. A detailed molecular belt model for apolipoprotein A-I in discoidal high density lipoprotein. J Biol Chem. 1999;274:31755–8.

35. Mei X and Atkinson D. Crystal structure of C-terminal truncated apolipoprotein A-I reveals the assembly of high density lipoprotein (HDL) by dimerization. J Biol Chem. 2011;286:38570–82.

36. Davidson WS and Thompson TB. The Structure of Apolipoprotein A-I in High Density Lipoproteins. J Biol Chem. 2007;282:22249–22253.

37. Thomas MJ, Bhat S and Sorci-Thomas MG. Three-dimensional models of HDL apoA-I: implications for its assembly and function. J Lipid Res. 2008;49:1875–83.

38. Lima DB, Melchior JT, Morris J, Barbosa VC, Chamot-Rooke J, Fioramonte M, Souza T, Fischer JSG, Gozzo FC, Carvalho PC and Davidson WS. Characterization of homodimer interfaces with cross-linking mass spectrometry and isotopically labeled proteins. Nat Protoc. 2018;13:431–458.

39. Forte T, Norum KR, Glomset JA and Nichols AV. Plasma lipoproteins in familial lecithin: cholesterol acyltransferase deficiency: structure of low and high density lipoproteins as revealed by elctron microscopy. J Clin Invest. 1971;50:1141–8.

40. Asztalos BF, Schaefer EJ, Horvath KV, Yamashita S, Miller M, Franceschini G and Calabresi L. Role of LCAT in HDL remodeling: investigation of LCAT deficiency states. J Lipid Res. 2007;48:592–9.

41. Rosenson RS, Brewer HB, Jr., Chapman MJ, Fazio S, Hussain MM, Kontush A, Krauss RM, Otvos JD, Remaley AT and Schaefer EJ. HDL measures, particle heterogeneity, proposed nomenclature, and relation to atherosclerotic cardiovascular events. Clin Chem. 2011;57:392–410.

42. Segrest JP, Jones MK, De Loof H, Brouillette CG, Venkatachalapathi YV and Anantharamaiah GM. The amphipathic helix in the exchangeable apolipoproteins: a review of secondary structure and function. J Lipid Res. 1992;33:141–66.

43. Shao B, Fu X, McDonald TO, Green PS, Uchida K, O’Brien KD, Oram JF and Heinecke JW. Acrolein impairs ATP binding cassette transporter A1-dependent cholesterol export from cells through site-specific modification of apolipoprotein A-I. J Biol Chem. 2005;280:36386–96.

44. Mei X, Liu M, Herscovitz H and Atkinson D. Probing the C-terminal domain of lipid-free apoA-I demonstrates the vital role of the H10B sequence repeat in HDL formation. J Lipid Res. 2016;57:1507–17.

45. Vedhachalam C, Ghering AB, Davidson WS, Lund-Katz S, Rothblat GH and Phillips MC. ABCA1-induced cell surface binding sites for ApoA-I. Arterioscler Thromb Vasc Biol. 2007;27:1603–9.

46. Phillips MC. Is ABCA1 a lipid transfer protein? J Lipid Res. 2018;59:749–763.

47. Segrest JP, Tang C, Song HD, Jones MK, Davidson WS, Aller SG and Heinecke JW. ABCA1 is an extracellular phospholipid translocase. Nat Commun. 2022;13:4812.

48. Heinecke JW. Small HDL promotes cholesterol efflux by the ABCA1 pathway in macrophages: implications for therapies targeted to HDL. Circ Res. 2015;116:1101–3.

49. Holleboom AG, Kuivenhoven JA, Vergeer M, Hovingh GK, van Miert JN, Wareham NJ, Kastelein JJ, Khaw KT and Boekholdt SM. Plasma levels of lecithin:cholesterol acyltransferase and risk of future coronary artery disease in apparently healthy men and women: a prospective case-control analysis nested in the EPIC-Norfolk population study. J Lipid Res. 2010;51:416–21.

50. Dullaart RP, Perton F, van der Klauw MM, Hillege HL, Sluiter WJ and Group PS. High plasma lecithin:cholesterol acyltransferase activity does not predict low incidence of cardiovascular events: possible attenuation of cardioprotection associated with high HDL cholesterol. Atherosclerosis. 2010;208:537–42.

