## Supplemental data for "Flipped C-Terminal Ends of APOA1 Promote ABCA1-dependent Cholesterol Efflux by Small HDLs"

### Supplemental Material

#### Supplemental Methods

**Materials**. Recombinant APOA1 (both regular and $^{15}N$-labeled) were generated as described.[21] The sequence of recombinant APOA1 was identical to that of mature human APOA1.

**Chemical crosslinking of APOA1 and isolation of r-HDL particles.** Reconstituted HDLs (r-HDLs) were crosslinked with EDC in phosphate-buffered saline (pH 6.5) and identified as described.[20,26] Crosslinking reactions were carried out in phosphate-buffered saline at 4°C with final concentrations of 20 mM EDC and 0.56 mg/mL APOA1 protein. After a 12-h incubation, the reactions were quenched with 1 M ammonium acetate (final concentration 50 mM). The reaction mixtures were further fractionated by high-resolution size exclusion chromatography (Superdex 200, 350 µL flow/min) to isolate monomeric HDL particles. Purified crosslinked HDL was exchanged into 50 mM ammonium bicarbonate with Amicon Ultra 10K centrifugal filter devices (Millipore, Eschborn, Germany) and stored at 4°C for MS/MS analysis.

**Proteolytic digestion for MS/MS analysis.** r-HDLs were incubated overnight at 37°C with sequencing grade modified trypsin (Promega, Madison, WI) at a ratio of 20:1 (w/w) protein/trypsin in 100 mM $NH_4HCO_3$, pH 8. Digestion was halted by acidification (pH 2–3) with trifluoroacetic acid.

**Tandem MS/MS analysis.** Capillary LC-ESI-MS/MS was performed using an IntegraFrit capillary trapping column packed with 1.5 cm of C18 (150 µM × 11 cm, New Objective, Woburn, MA; Magic C18, 5 µM, 200 Å, Michrom BioResources, Auburn, CA), a capillary analytical column packed with 15 cm of C18 (75 µM × 15 cm, Magic C18, 5 µM, 100 Å, Michrom BioResources, Auburn, CA), an LTQ Orbitrap XL mass spectrometer (Thermo Electron, Bremen, Germany), and a nanoACQUITY UPLC system (Waters, Milford, MA). To remove salts and contaminants in each experiment, ~1 µg of tryptic digest was injected onto the trapping column and flushed for 10 min with a mobile phase of 0.1% TFA in 97:3 water/ACN at a flow rate of 3 µL/min. The flow rate was then reduced to 0.3 µL/min, effluent from the trapping column was directed to the capillary LC column, and an 80-min gradient between 2% and 40% mobile phase B (0.1% TFA in ACN) was implemented. Eluting peptides were electrosprayed into an LTQ-Orbitrap mass spectrometer operating in data-dependent mode to acquire a full MS scan (400–2000 m/z), and subsequent MS/MS scans were acquired for the five most intense precursor ions. CID of the precursors occurred in the LTQ at 35% normalized collision energy. Isolation width was set to 2 m/z, and monoisotopic precursor selection was enabled. MS/MS scans were acquired in both the ion trap and orbitrap. Charge state rejection was enabled for 1+ and 2+ charge states, as crosslinked peptides formed in these experiments tend to have a higher charge state upon ESI than non-crosslinked (linear peptide) species. Rejection of low charge states from MS/MS acquisition provides a useful bias for detecting crosslinks. MS/MS spectrum lists in the raw files were extracted and converted to mzXML files, using ReAdW (version 4.6.0) for database searching.

**MS data analysis.** The MS/MS data in the mzXML files were subsequently searched against the database for crosslinks, using xQuest (version 2.1.1).[26] The database is built from the sequence of APOA1 for the r-HDL experiment or 39 HDL proteins for human

HDL crosslinking results. xQuest is an algorithm that can search for theoretical crosslinks whose masses match measured precursor masses and to subsequently assign the fragment masses of MS/ MS spectra. In its interpretation of MS/MS spectra, xQuest assumes that a crosslink precursor will fragment at only one peptide bond. For our searches, xQuest was used with default settings, except that mass shifts for crosslinking products were manually set to −18.010564686 for the “xlink mass-shift.”

The modified residues were set to Lys, Asp, and Glu. Matches from all searches were required to have precursor mass errors ≤4.3 ppm and ≥5% of the ion current in a given MS/MS spectrum assigned as b- and y-type ions. xQuest matches for intra-protein and inter-protein crosslinks were required to meet different scoring thresholds. Intra-protein crosslink matches were required to have xQuest scores ≥25. Inter-protein crosslink matches were required to have xQuest scores ≥29. Inter-protein crosslink matches were disregarded unless three unique fragments were assigned on each peptide chain. Spectra matches were assessed for this characteristic after assigning neutral losses and second isotopic peaks. In addition, crosslink matches were required to have at least 8 of these sequence-specific fragments assigned. Finally, spectra that corresponded to crosslinks, either intra-protein or inter-protein, with multiple possible linkage patterns, were manually inspected. Exact linkages were proposed only when two crosslinked residues could be unequivocally defined by the observed fragmentation.

**Incubation of control and LCAT-deficient plasma with LCAT*.*** Plasma (150 µL) from 3 LCAT-/- subjects and 3 control subjects were incubated with or without 50 µg/mL recombinant human LCAT[32] at 37ºC for 1 h followed by addition of DTNB (2 mM final concentration) to inhibit the enzyme. Because DTNB interferes with Amplex red cholesterol assay, prior to addition of DTNB an aliquot (20 µL) was taken for cholesterol assays and immediately chilled on ice. ABCA1-specific CEC was measured in ABCA1-expressing BHK cells as described in Methods, **Figure 5** and **Supplemental Fig. S3**.

**Molecular dynamics (MD) simulations of the APOA1 structures in rHDL.** We used Anton 2 (a specialized supercomputer designed to accelerate molecular dynamics simulations) to study the dynamic structure of APOA1 molecules. Anton 2 rapidly generates all-atom trajectories for much longer time intervals (10’s of microseconds) than possible on other computer systems which greatly increases confidence in the results because of increased sampling.

**All-atom and CG simulations of r-HDL-90 (20 µs all-atom, 15 µs simulated tempering, 200 µs CG).** Structures for r-HDL-80 and r-HDL-90 were generated using a sequence of conventional all-atom MD, simulated tempering (ST), and coarse-grained (CG) simulations.[27-29] The initial configuration of a 20 µs-long all-atom simulation of r-HDL-90 was developed by removing POPC and cholesterol from a disc of r-HDL-100.[27] The disc was solvated by water and 0.15 mM NaCl, using CHARMM-GUI.[29] The box was cubic ($A = B = C = 137$ Å) and the pressure was applied isotropically. The total number of particles was ~258,000, including 100 POPC and 10 cholesterol. The system was first equilibrated for ~2 ns on an in-house computer cluster (Biowulf) to generate an initial structure that was then further developed on the Anton-2 supercomputer.

To enhance sampling of the conformation space, a series of successive ST simulations totaling 15 µs on the Anton-2 supercomputer extended the 20 µs-long conventional MD simulation of r-HDL-90. One hundred and seventeen temperatures, ranging from 310 K to 450 K and distributed exponentially,[28] comprised the temperature ladder of ST simulations. Prior to ST simulations, the 20 μs frame of conventional MD was simulated for 5 ns at the 117 temperatures. The following equation determined the weights for the first ST simulation:

$$g_{n+1} - g_n = \left(\frac{1}{k_B T_{n+1}} - \frac{1}{k_B T_n}\right)\frac{E_{n+1}+E_n}{2} \quad , \qquad \textit{Equation 1}$$

where $g$, $T$, $E$, and $k_B$ are the weight, temperature, energy, and Boltzmann's constant. The weights and maximum temperatures were further adjusted for subsequent ST simulations on an ad hoc basis to allow for more frequent transitions and to avoid long residence at certain temperatures. The exchange was allowed between adjacent temperature ladders with the following probability:

$$p(T_n \rightarrow T_{n+1}) = min\left\{1\,, e^{\left(\frac{1}{k_B T_n} - \frac{1}{k_B T_{n+1}}\right)E_n + (g_{n+1} - g_n)}\right\}. \qquad \textit{Equation 2}$$

Note that the probability is determined by the weight difference between two ladders rather than each weight.

A 200 μs-long CG simulation with the Martini force field on Gromacs (www.gromacs.org, version 2016.6) completed the study of r-HDL-90. The CG simulation frames were converted to all-atom for developing the contact maps.

**All-atom and CG simulations of r-HDL-80 (1 μs all-atom, 200 μs CG).** The initial configuration of a 1 μs-long all-atom simulation of r-HDL-80 was developed by removing POPC and cholesterol from the last configuration of the 20 μs-long conventional MD of r-HDL-90. The disc was solvated by water and 0.15 mM NaCl, using CHARMM-GUI.[29] The number of phospholipid and cholesterol molecules in each size of r-HDL were determined using nondenaturing gradient gel electrophoresis.[30] The box was cubic ($A = B = C = 126$ Å) and the pressure was applied isotropically. The total number of particles was ~ 200,000, including 50 POPC and 5 cholesterol (the number of phospholipid and cholesterol molecules are per particle, following the notation of our previous simulations.[27] We choose the relatively large box size for r-HDL-80 (compared to that of r-HDL-90) because its APOA1 terminals exhibit enhanced mobility relative to r-HDL-90 particles. The use of a sufficiently large box size ensured that the protein does not interact with its periodic image during MD simulation. Consequently, reducing the number of lipids in rHDL-90 by half to obtain rHDL-80 does not necessarily result in a significantly smaller box size.

The system was first analyzed for ~ 2 ns on an in-house computer cluster (Biowulf). The main simulation was carried out on the Anton-2 supercomputer. A 200 μs-long CG simulation with the Martini force field on Gromacs completed the study of r-HDL-80. The CG simulation frames were converted to all-atom for developing the contact maps.

**Additional details of all-atom simulations.**
CHARMM36 was used for lipid and protein force field parameters. The TIP3P water model modified for CHARMM was used to describe water molecules. Lennard-Jones (LJ) parameters of $Na^+$ and $Cl^-$ as well as $Na^+$ and selected oxygens of lipids and proteins were taken from the CHARMM 36 ion force field parameters (NBFIX).

In-house all-atom trajectories were generated using CHARMM with a leapfrog Verlet algorithm and a time step of 2 fs. Temperature and pressure were kept at 310 K and 1 bar using Nose-Hoover thermostat and Langevin piston barostat ($\gamma = 0$), respectively. Masses of the temperature and pressure pistons were 20% and 2% of the system masses, respectively. Lennard–Jones potentials were terminated at 12 Å, with a smoothing function operating between 8 Å and 12 Å. Electrostatics were evaluated using particle-mesh Ewald with approximately 1 grid point per Å, a sixth-order spline interpolation for the complementary error function, a real-space cutoff of 12 Å, and $k$ = 0.32. All bonds to hydrogen atoms were constrained using SHAKE.

For the Anton2 simulation, a multigrator, which minimizes sources of error associated with limited-precision arithmetic and truncation errors, generated trajectories with a time step of 2 fs. Temperature and pressure were kept constant at 310 K and 1 bar, respectively, using a variant of Nosé–Hoover and the Martyna–Tobias–Klein. Electrostatic forces were calculated using the u-series method. Water molecules and all bond lengths to hydrogen atoms were constrained using M-SHAKE.

#### Supplemental Tables

**Table S1. LCAT mutations (nomenclature according to HGMD) of the Italian cohort**[31]

| **Nucleotide Change** | **Amino acid change** | **Genotype** | **Number of carriers** |
|---|---|---|---|
| c.511C>T | Arg171Trp | Homozygote | 1 |
| c.893C>T | Thr298Ile | Homozygote | 1 |
| c.1007A>G/c.1132G>A | Tyr336Cys/Glu378Lys | Compound | 2 |
| c.31delG | Val11* | Heterozygote | 2 |
| c.511C>T | Arg171Trp | Heterozygote | 1 |
| c.893C>T | Thr298Ile | Heterozygote | 3 |

**Table S2. Crosslinks in r-HDLs identified by MS/MS analysis.**

| | r-HDL-80 | | | r-HDL-88 | | | r-HDL-96 | | | r-HDL-120 | | |
|---|---|---|---|---|---|---|---|---|---|---|---|---|
| | Crosslinks | XL Type | Relative Intensity | Crosslinks | XL Type | Relative Intensity | Crosslinks | XL Type | Relative Intensity | Crosslinks | XL Type | Relative Intensity |
| N-terminus | K23-D51 | intra | Weak | K23-D28 | intra | Weak | K23-D48 | intra | Medium | D28-K133 | intra | Weak |
| | D28-K12 | intra | Weak | E34-K45 | intra | Strong | K45-E34 | intra | Medium | E80-K88 | intra | Medium |
| | K45-E34 | intra | Strong | E76-K12 | intra | Weak | E76-K12 | intra | Medium | | | |
| | K59-D28 | intra | Weak | | | | | | | | | |
| | K59-E78 | intra | Weak | | | | | | | | | |
| | D73-K12 | intra | Weak | | | | | | | | | |
| | E76-K12 | intra | Weak | | | | | | | | | |
| Center region | K96-E169 | inter | Strong | K96-E169 | inter | Strong | K96-E147 | inter | Medium | K96-E169 | inter | Strong |
| | K96-E147 | inter | Weak | K96-E146 | inter | Weak | K96-D168 | inter | Medium | K96-D168 | inter | Weak |
| | K106-E125 | inter | Weak | K140-E125 | inter | Medium | K96-E169 | inter | Strong | K96-E146 | inter | Weak |
| | K140-E125 | inter | Weak | K195-E70 | inter | Strong | K107-E125 | inter | Weak | K118-E147 | inter | Medium |
| | | | | | | | K107-D157 | inter | Medium | E125-K118 | inter | Weak |
| | | | | | | | K118-E147 | inter | Strong | K133-E128 | inter | Weak |
| | | | | | | | K133-E111 | inter | Weak | K133-E125 | inter | Medium |

| | | | | | | | | | | | | |
|---|---|---|---|---|---|---|---|---|---|---|---|---|
| | | | | | | | K133-E125 | inter | Medium | E139-K118 | intra | Weak |
| | | | | | | | K140-E125 | inter | Strong | K140-E125 | inter | Strong |
| | | | | | | | | | | E147-K133 | intra | Weak |
| C-terminus | K77-E191 | inter | Medium | K77-E191 | inter | Medium | K195-E70 | inter | Medium | K182-E191 | intra | Weak |
| | K96-E198 | intra | Weak | K96-D198 | inter | Weak | K208-E198 | intra | Medium | E198-K208 | intra | Weak |
| | K96-E235 | intra | Medium | K195-E78 | inter | Medium | K208-E223 | intra | Medium | E205-K208 | intra | Strong |
| | K195-E234 | intra | Medium | K195-E235 | intra | Weak | | | | E198-K182 | intra | Weak |
| | K226-E78 | intra | Weak | E234-K208 | intra | Weak | | | | | | |
| | K226-E205 | intra | Weak | E234-K182 | intra | Medium | | | | | | |
| | E235-K182 | intra | Medium | E235-K208 | intra | Weak | | | | | | |
| N-C terminus links | D48-K238 | inter | Weak | | | | K238-E34 | intra | Weak | | | |

**Table S3. HDL particle concentration of study subjects as determined by calibrated IMA.**

| HDL Species | Control subjects (n=14) | LCAT-heterozygous subjects (n=6) | LCAT-deficient subjects (n=6) | *P*-value |
|---|---|---|---|---|
| Total-HDL (μM) | 14.9±2.5 | 8.3±2.9 | 2.9±1.3 | <0.0001 |
| XS-HDL (μM) | 0.6±0.4 | 1.6±0.7 | 1.2±0.7 | 0.0017 |
| S-HDL (μM) | 1.9±1.2 | 2.5±1.0 | 0.4±0.6 | 0.009 |
| M-HDL (μM) | 7.7±1.8 | 3.4±1.4 | 0.5±0.5 | <0.0001 |
| L-HDL (μM) | 4.6±1.7 | 0.8±0.4 | 0.9±0.5 | <0.0001 |

*P*-values are from a mixed effect model and Tukey-Kramer post-tests.

#### Supplemental Figures

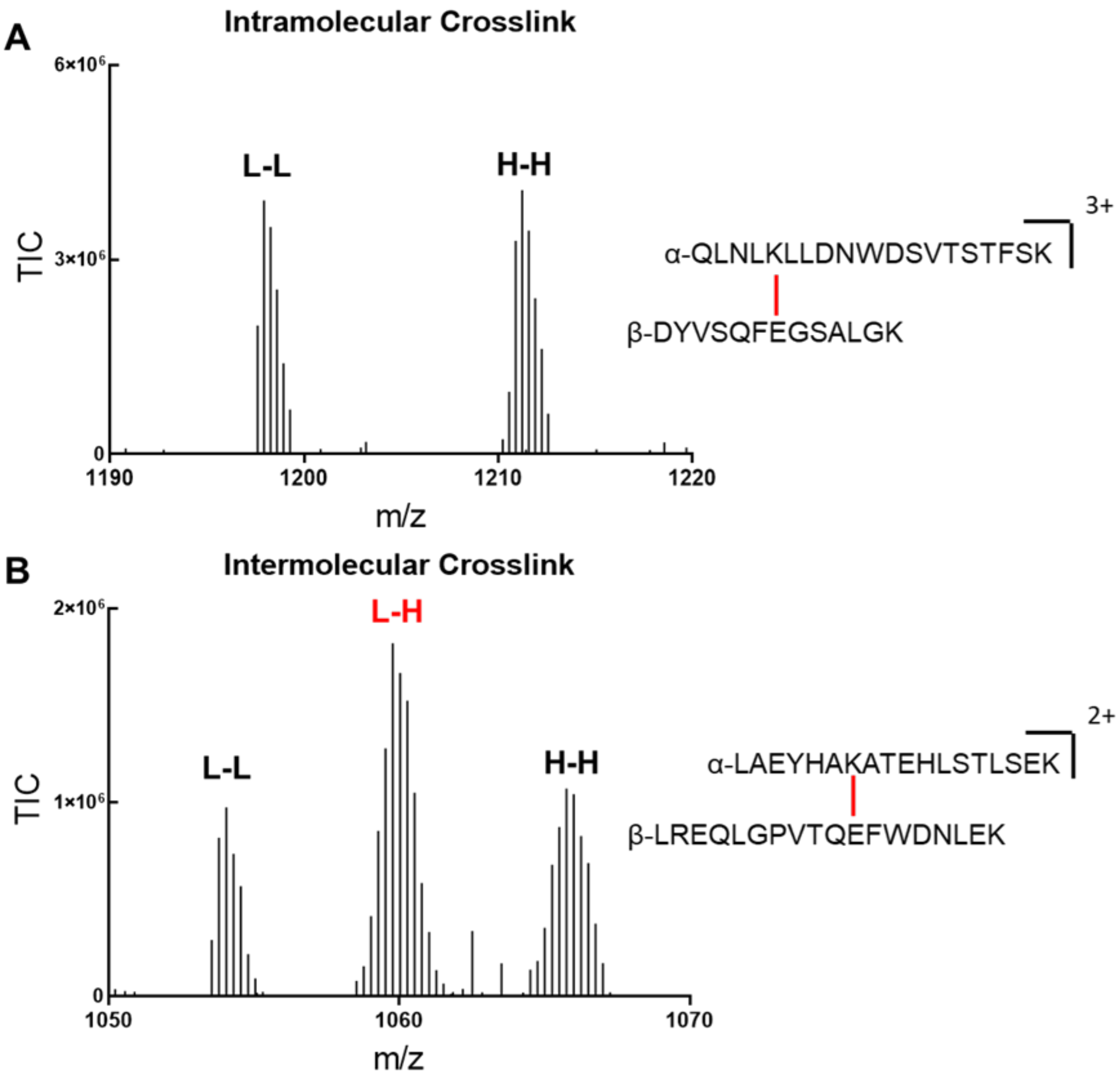


**Figure S1. Representative MS spectra of an (A) intramolecular crosslink (K45–E34) and (B) intermolecular crosslink (K195–E70) of APOA1 detected in r-HDL**. MS1 spectra of (A) intra-protein crosslink QLNL**K**LLDNWDSVTSTFSK-DYVSQF**E**GSALGK and (B) inter-protein crosslink LAEYHA**K**ATEHLSTLSEK-LREQLGPVTQ**E**FWDNLEK.

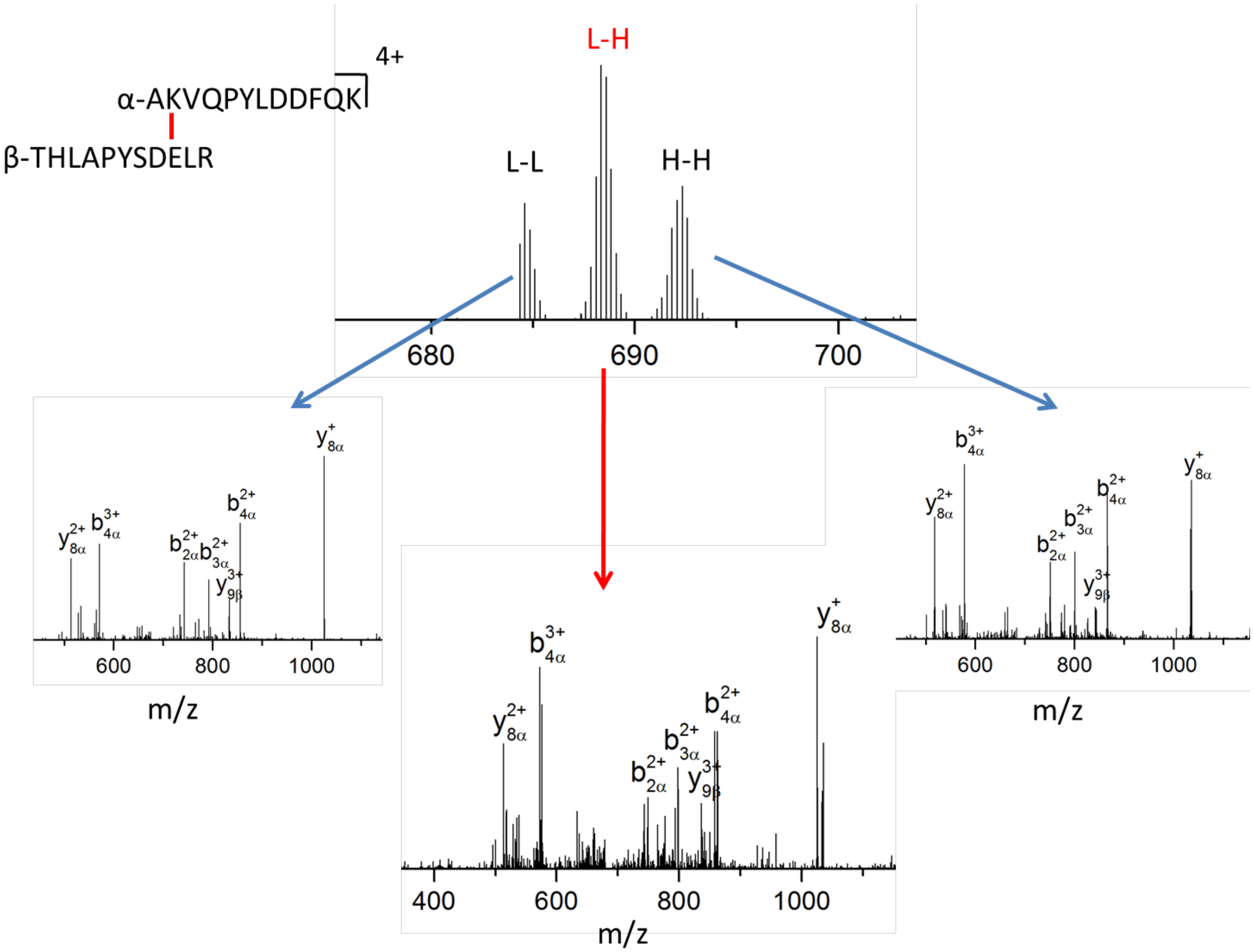


**Figure S2. Representative MS1 and MS/MS spectra of the intermolecular crosslink K96–E169 of APOA1 in r-HDL**. Interpeptide crosslink AKVQPYLDDFQK-THLAPYSDELR.

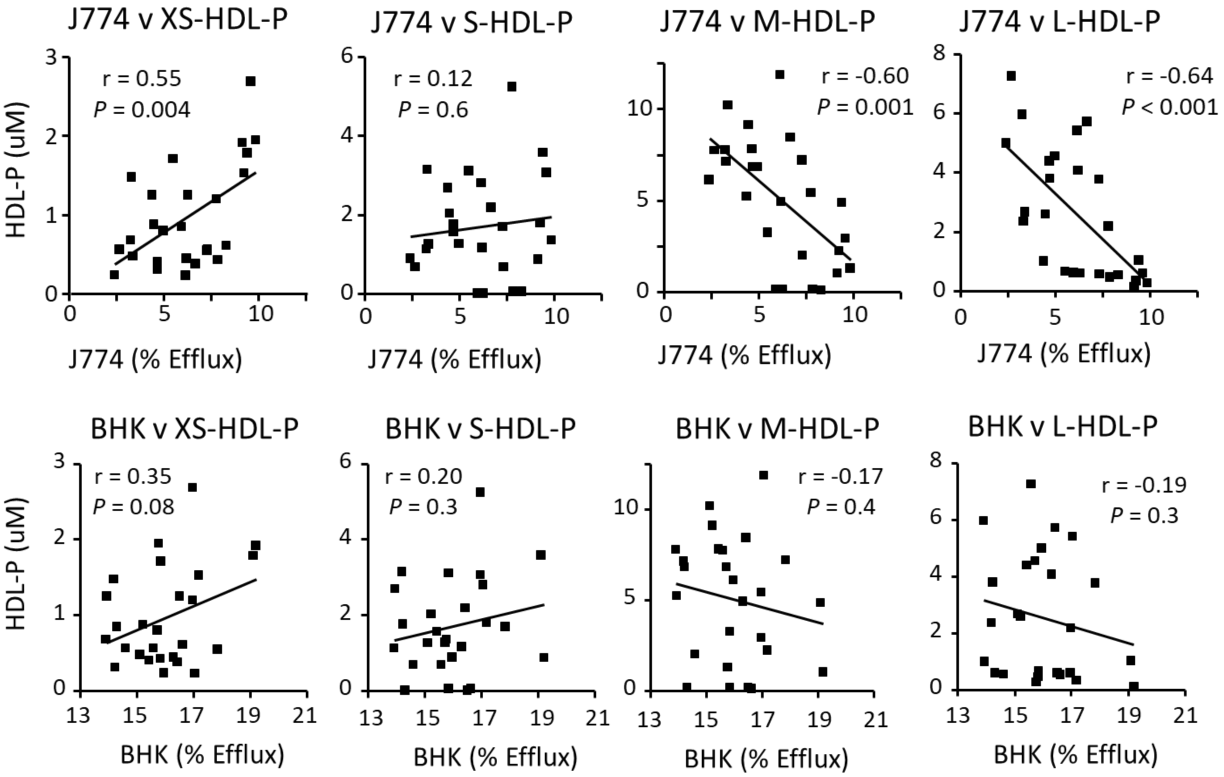


**Figure S3. Scatter plots of the correlations of macrophage (J774 cells) and ABCA1 (BHK cells) CEC of serum HDL with HDL subspecies for all subjects in the LCAT study cohort**. CEC was quantified as described in Fig. 5 of the paper. r, Pearson correlation.
